# Validation of SeptiCyte RAPID to discriminate sepsis from non-infectious systemic inflammation

**DOI:** 10.1101/2022.07.20.22277648

**Authors:** Robert Balk, Annette M. Esper, Greg S. Martin, Russell R. Miller, Bert K. Lopansri, John P. Burke, Mitchell Levy, Steven Opal, Richard E. Rothman, Franco R. D’Alessio, Venkataramana K. Sidhaye, Neil R. Aggarwal, Jared A. Greenberg, Mark Yoder, Gourang Patel, Emily Gilbert, Jorge P. Parada, Majid Afshar, Jordan A. Kempker, Tom van der Poll, Marcus J. Schultz, Brendon P. Scicluna, Peter M.C. Klein Klouwenberg, Janice Liebler, Emily Blodget, Santhi Kumar, Krupa Navalkar, Thomas D. Yager, Dayle Sampson, James T. Kirk, Silvia Cermelli, Roy F. Davis, Richard B. Brandon

**Affiliations:** Rush Medical College and Rush University Medical Center, Chicago, Illinois; Grady Memorial Hospital and Emory University School of Medicine, Atlanta; FirstHealth of the Carolinas, Pinehurst, North Carolina; Intermountain Medical Center, Murray, Utah; University of Utah School of Medicine, Salt Lake City, Utah; Brown University, Providence, Rhode Island; Johns Hopkins University School of Medicine, Baltimore, Maryland; Anschutz Medical Campus, University of Colorado, Denver, Colorado; Loyola University Medical Center, Maywood, Illinois; University of Wisconsin School of Medicine and Public Health, Madison, Wisconsin; Amsterdam UMC, University of Amsterdam, Amsterdam, the Netherlands; Centre for Molecular Medicine and Biobanking, University of Malta, Malta; and Department of Applied Biomedical Science, Faculty of Health Sciences, Mater Dei hospital, University of Malta, Malta; Isala Hospital, Zwolle, the Netherlands; Keck Hospital of University of Southern California (USC); and Los Angeles County and USC Medical Center; Immunexpress Inc., Seattle, Washington

**Keywords:** sepsis, diagnosis, host response, SIRS, sepsis scoring systems

## Abstract

**(1) Background:** SeptiCyte RAPID is a molecular test for discriminating sepsis from non-infectious systemic inflammation, and for estimating sepsis probabilities. The objective of this study was the clinical validation of SeptiCyte RAPID, based on testing retrospective banked and prospectively collected patient samples.

**(2) Methods:** The cartridge-based SeptiCyte RAPID test accepts a PAXgene blood RNA sample and provides sample-to-answer processing in ∼1 hour. The test output (SeptiScore, range 0-15) falls into four interpretation bands, with higher scores indicating higher probabilities of sepsis. Retrospective (N=356) and prospective (N=63) samples were tested from adult patients in ICU either having the systemic inflammatory response syndrome (SIRS), or suspected of / diagnosed with sepsis. Patients were clinically evaluated by a panel of three expert physicians blinded to the SeptiCyte test results. Results were interpreted under either the Sepsis-2 or Sepsis-3 framework.

**(3) Results:** Under the Sepsis-2 framework, SeptiCyte RAPID performance for the combined retrospective and prospective cohorts had Area Under the ROC Curve (AUC) ranging from 0.82 to 0.85, negative predictive value 0.91 (sensitivity 0.94) for SeptiScore Band 1 (score range 0.1-5.0; lowest risk of sepsis), and positive predictive value 0.81 (specificity 0.90) for SeptiScore Band 4 (score range 7.4-15; highest risk of sepsis). Performance estimates for the prospective cohort ranged from AUC 0.86-0.95. For physician-adjudicated sepsis cases that were blood culture (+) or blood, urine culture (+)(+), 43/48 (90%) of SeptiCyte scores fell in Bands 3 or 4. In multivariable analysis with up to 14 additional clinical variables, SeptiScore was the most important variable for sepsis diagnosis. Comparable performance was obtained for the majority of patients reanalyzed under the Sepsis-3 definition, although a subgroup of 16 patients was identified that was called septic under Sepsis-2 but not under Sepsis-3.

**(4) Conclusions:** This study validates SeptiCyte RAPID for estimating sepsis probability, under both the Sepsis-2 and Sepsis-3 frameworks, for hospitalized patients on their first day of ICU admission.

## 1. Introduction

Sepsis is an important and expensive global health problem with high morbidity and mortality. According to the World Health Organization, more than 11 million people die from sepsis worldwide annually - more than the deaths caused by all cancers combined [2] and comprising almost 20% of all deaths annually [1]. Sepsis is the leading cause of death in U.S. hospitals and is ranked as the most expensive disease state to manage for admitted patients [3], with total annual costs of treatment and rehabilitation estimated at $62 billion [4]. Before the COVID-19 pandemic, at least 1.7 million adults in the United States developed sepsis annually [5]. During the pandemic, this problem was exacerbated as a large fraction of COVID-19 deaths appeared due to viral and/or bacterial sepsis [6, 7].

Early identification of sepsis and implementation of treatment bundles have been shown to improve outcomes for sepsis patients [8]. However, early identification of sepsis is difficult for many reasons. In the early stages of the disease, patients present with inflammatory clinical signs which may be early indicators of a septic response, but which are also common to many other non-infectious conditions [9]. Proof of infection by the conventional ‘gold standard’ criterion of culture positivity lacks timeliness [10] and cultures are negative in a significant fraction of retrospectively diagnosed sepsis cases [11,12]. The presence of infection could perhaps be inferred from the clinician’s act of therapeutic antibiotic administration, but this is also problematic, as antibiotics are found in retrospect to often be overprescribed [13–15]. Also, in the early stages of sepsis, organ dysfunction (the *<u>sine qua non</u>* for sepsis under the Sepsis-3 definition) may not yet be highly evident or easily detected. In short, the detection of sepsis in its early stages where treatment could be most effective (i.e. before organ damage is extensive) remains a challenge. This point of concern has been raised by a number of authors in connection with the Sepsis-3 definition [16–21].

We previously reported on the development and validation of a diagnostic test (SeptiCyte LAB) to differentiate patients with sepsis from those exhibiting a non-infectious systemic inflammatory response syndrome (SIRS). The test provided a probability of sepsis, based on measurement of four host immune response biomarkers, PLA2G7, PLAC8, CEACAM4 and LAMP1 [22,23]. Here we describe SeptiCyte RAPID, a simplified and improved cartridge-based version of the earlier SeptiCyte LAB test, which achieves simultaneous amplification and detection of two of the original four RNA transcripts (PLA2G7 and PLAC8) in human blood samples. The specific aim of the present work is to provide clinical validation data demonstrating robust performance of SeptiCyte RAPID for the estimation of sepsis probabilities, under both the Sepsis-2 and Sepsis-3 frameworks, in a combined cohort of 419 hospitalized patients on their first day of ICU admission. SeptiCyte RAPID addresses a clinical need for more rapid and accurate differentiation of sepsis from SIRS within a clinically actionable (∼1 hour) time frame. Some of these results have been presented earlier in the form of an abstract [24].

## 2. Materials and Methods

### 2.1 Study cohorts

Clinical validation of SeptiCyte RAPID for the discrimination of sepsis from SIRS according to the Sepsis-2 definition used PAXgene blood RNA samples from retrospective (N=356) and prospective (N=63) patient cohorts. A flow diagram describing the origin of all samples used in the study is provided in the Supplementary Information, Section 1.

The retrospective cohort was drawn from the observational MARS, VENUS and VENUS Supplement trials (www.clinicaltrials.gov NCT01905033 and NCT02127502) which have been previously described [23]. The retrospective cohort comprised 80% of the 447 patients used for the 510(k) clearance of SeptiCyte LAB, for which duplicate banked PAXgene blood RNA samples remained available. The recruitment dates were Jan 2011 - Dec 2013 (MARS), May 2014 - Apr 2015 (VENUS) and Mar - Aug 2016 (VENUS Supplement).

The prospective cohort consisted of 63 critically ill adult subjects enrolled in an observational trial (NEPTUNE, www.clinicaltrials.gov NCT05469048) between the dates May 26, 2020 - April 25, 2021 at Emory University / Grady Memorial Hospital (Atlanta GA), Rush University Medical Center (Chicago IL) and University of Southern California (USC) Medical Center (with two separate sites, Keck Hospital of USC, and Los Angeles county + USC Medical Center, in Los Angeles CA). By design, the NEPTUNE inclusion/exclusion criteria matched the criteria used earlier in the MARS, VENUS, and VENUS Supplement studies. Subjects were considered for inclusion in NEPTUNE if they were adults (≥18 years old), exhibited two or more SIRS criteria, and received an ICU admission order. Subjects were operationally defined as “suspected of sepsis” if microbiological tests were ordered within 24 hours of the ICU admission order. Subjects were excluded if therapeutic intravenous antibiotic treatment was initiated >24 hours before ICU admission order, as this would be expected to decrease blood culture positivity [25,26], thus potentially confounding the retrospective physician diagnosis (RPD) process. Extensive early antibiotic treatment could also potentially affect the host response to infection, introducing bias into the SeptiCyte RAPID test scores. As much as possible, enrolments were consecutive; however, during the COVID-19 pandemic, screening did not happen every day, and initial consenting sometimes resulted in later refusal to participate. PAXgene blood samples were collected within 24 hours of ICU admission order and run fresh on Idylla instruments installed at the sites. Clinical data were collected on case report forms as described previously [23].

### 2.2 Ethics approval and consent to participate

Ethics approval for the MARS trial was given by the Medical Ethics Committee of the Amsterdam Medical Center (approval # 10-056C). Ethics approvals for the VENUS trial were given by the relevant Institutional Review Boards as follows: Intermountain Medical Center/Latter Day Saints Hospital (approval # 1024931); Johns Hopkins Hospital (approval # IRB00087839); Rush University Medical Center (approval # 15111104-IRB01); Loyola University Medical Center (approval # 208291); Northwell Healthcare (approval #16-02-42-03). Ethics approvals for the NEPTUNE trial were given by the relevant Institutional Review Boards as follows: Emory University (approval # IRB00115400); Grady Memorial Hospital (approval # 00-115400); Rush University Medical Center (approval # 19101603-IRB01); University of Southern California Medical Center (approval # HS-19-0884-CR001). All subjects, or their legally authorized representatives, gave informed consent for participation in this study. All methods used in this study were carried out in accordance with the relevant guidelines and regulations.

### 2.3 The SeptiCyte RAPID test

SeptiCyte RAPID is run on the Idylla platform (Biocartis NV, Mechelen, Belgium). The test is performed by pipetting 0.9 mL of PAXgene-stabilized blood (corresponding to 0.24 mL of drawn blood) into a custom cartridge which performs all assay steps including sample extraction/purification and RT-qPCR for the detection and relative quantification of the PLAC8 and PLA2G7 mRNA targets. Test results are calculated and presented automatically through a software-generated report which includes a quantitative score (SeptiScore, range 0-15), calculated as the difference between the RT-qPCR Cq values for PLA2G7 and PLAC8 and proportional to sepsis probability. The test has a hands-on time of ∼2 min and a turnaround time of ∼1 hour.

### 2.4 Reference diagnosis (comparator) under Sepsis-2

Clinical performance of SeptiCyte RAPID was evaluated by comparison to a ‘gold standard’ of clinical adjudication. The adjudication process, termed retrospective physician diagnosis (RPD), consisted of clinical evaluations by an external three-member panel of experienced physicians not involved in the care of the patients. For each patient, a chart review was conducted independently by each panel member, leading to a three-way classification (sepsis, SIRS or indeterminate) in accordance with the Sepsis-2 definition under which sepsis is defined as ≥2 SIRS criteria + infection [27]. The vote tally for each patient was either unanimous, consensus (2 of 3), or indeterminate. There were 41/419 (9.8%) indeterminates, which were then reanalyzed by forcing into either the sepsis or SIRS categories by a second RPD. The Sepsis-2 definition was used in this initial analysis because the majority of patients were recruited before the Sepsis-3 definition became available. Additional details are provided in Supplementary Information, Section 2.

### 2.5 Reference diagnosis (comparator) under Sepsis-3

An additional analysis was conducted under the Sepsis-3 definition. Figure 1 of Singer et al. (2016) [28] describes how we operationalized the Sepsis-3 definition. Singer et al. proposed that patients with organ dysfunction (SOFA scores ≥2) and with definite or suspected infection could be considered to have sepsis. Therefore, the key to transitioning from a Sepsis-2 to Sepsis-3 framework lies in determining whether patients with organ dysfunction (SOFA scores ≥2) are infection-positive or infection-negative. Under the Sepsis-2 framework, when the RPD panelists called a patient septic, this implied that the patient exhibited ≥2 SIRS criteria *<u>and</u>* had a definite or probable infection. Therefore, a call of sepsis by the RPD panelists implied that they classified the patient as infection-positive.

**Figure 1.**
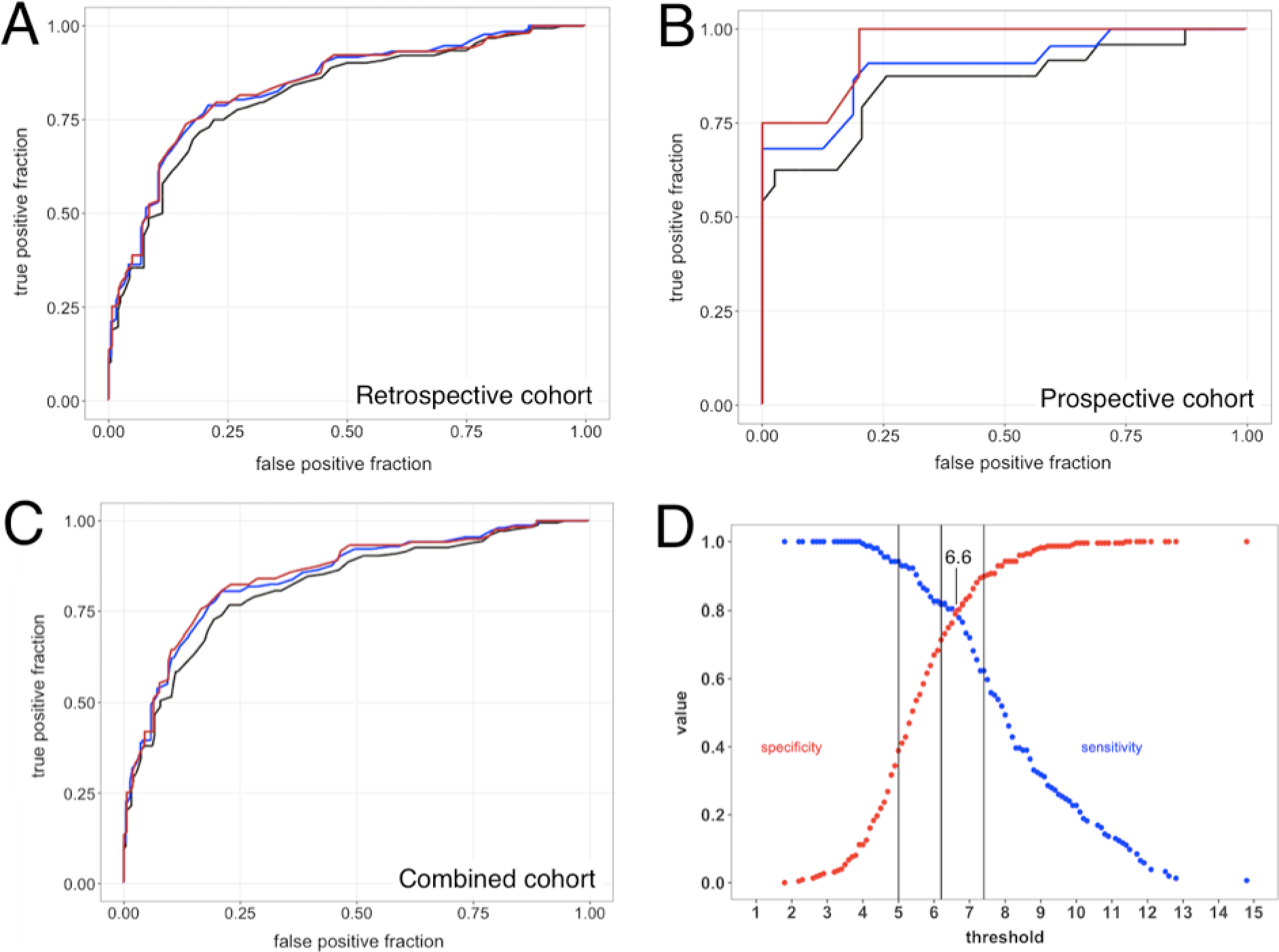
Diagnostic performance of SeptiCyte RAPID. ROC curve analyses for SeptiCyte RAPID were performed using three different RPD methods (consensus, forced and unanimous), for either the retrospective, prospective, or full cohorts. **(A)** Retrospective cohort ROC curves for forced (black), consensus (blue) and unanimous (red) RPD, with AUC = 0.82, 0.85 and 0.85 respectively. **(B)** Prospective cohort ROC curves for forced (black), consensus (blue) and unanimous (red) RPD, with AUC = 0.86, 0.90 and 0.95 respectively. **(C)** Combined cohort ROC curves for forced (black), consensus (blue) and unanimous (red) RPD, with AUC = 0.82, 0.85 and 0.85 respectively. **(D)** Sensitivity (blue) and specificity (red) as a function of threshold SeptiScore. Comparator = consensus RPD. Band boundaries at 5.0, 6.2, 7.4 are shown as vertical black lines.

As measures of organ dysfunction, we considered both the complete SOFA score [29] which had been collected for 289/419 (69.0%) of patients, and also, following Grissom et al. [30] and Lambden et al. [31], a partial SOFA score (pSOFA) in which 4 or more of the 7 SOFA components were present. The pSOFA could be computed for 372/419 (88.8%) of patients.

### 2.6 Statistical calculations

Sepsis probability as a function of SeptiScore was calculated by a “sliding window” approach [32]. A custom R script was written to conduct a sliding window calculation of the sepsis probability (P) across the 0-15 SeptiScore range. The sliding window was 4 SeptiScore units wide and was shifted in 1-unit increments from the lower to the upper limit of the SeptiScore range. In each 4-unit wide window, the numbers of sepsis calls (N_sepsis_) and SIRS calls (N_SIRS_) were tabulated according to each RPD method, and P = (N_sepsis_) / (N_sepsis_ + N_SIRS_) was computed for that window. The process was repeated stepwise over the entire 0-15 SeptiScore range.

Sepsis probability as a function of SeptiScore was parsed into four bands, with higher SeptiScores representing higher probabilities of sepsis. Band boundaries were pre-defined on the basis of an independent set of 195 clinical samples from the MARS consortium. The samples for setting the band boundaries were independent of the 419 samples in the validation dataset, and were not used in the performance evaluation. The band boundaries were set to give 90% sensitivity for binarization at the Band 1/2 boundary (SeptiScore 4.95) and 80% specificity for binarization at the Band 3/4 boundary (SeptiScore 7.45), using site clinical adjudications as Ground Truth values. The intermediate zone between the Band 1/2 and Band 3/4 boundaries was divided in half to define the Band 2/3 boundary (SeptiScore 6.15).

Receiver-Operating Characteristic (ROC) curve analysis was performed in accordance with Clinical and Laboratory Standards Institute EP24-A2 using the pROC package [33]. Other clinical performance measures (clinical sensitivity, clinical specificity, sepsis and SIRS probabilities, and likelihood ratios) were also calculated. Additional statistical methods for data analyses, including methods for imputing missing data and for combining the SeptiScore with other clinical variables, are described in the Supplementary Information, Sections 3-5.

## 3. Results

### 3.1 Description of the SeptiCyte RAPID test

SeptiCyte RAPID is a fully automated sample-to-result test, with all reagents integrated in a single-use cartridge. The test provides an actionable result in ∼1 hour, in the form of a SeptiScore ranging from 0 to 15 and falling into one of four “SeptiScore Bands” of increasing sepsis probability. SeptiCyte RAPID provides a significant technical advance over the earlier SeptiCyte LAB assay which was based upon the conventional 96-well Applied Biosystems 7500 Fast Dx format with a turnaround time of ∼8 hours [23]. Correlation of SeptiScore values between SeptiCyte LAB and SeptiCyte RAPID, based upon the retrospective sample set (N=356) run on both platforms, was high with Pearson’s sample correlation coefficient r = 0.88. (Supplementary Information, Section 6).

SeptiCyte RAPID quantitatively measures the relative expression levels of two host immune response genes PLAC8 (Placenta Associated 8) and PLA2G7 (Phospholipase A2 Group 7). This expression signature was discovered using a purely bioinformatic approach which compared SIRS patients that had been retrospectively diagnosed as having either infection or no infection [22]. With respect to biological roles in the host immune response to infection, PLAC8 is reportedly an interferon inducible gene expressed in a variety of tissues rich in immune cells (e.g. whole blood, spleen, lymph node, colon), and up-regulated in sepsis across a broad range of different peripheral blood cell types including plasmacytoid dendritic and natural killer cells. PLA2G7 encodes the protein platelet-activating factor (PAF) acetylhydrolase, a secreted enzyme primarily produced by macrophages that catalyzes the degradation of PAF and hydrolyses the oxidized short chain phospholipids of low-density lipoproteins (LDL), thereby releasing pro-inflammatory mediators (lysophospholipids and oxidized fatty acids). Further discussion and literature references are provided in the Supplementary Information, Section 7.

### 3.2 Demographic and clinical characteristics of the study cohorts

Clinical validation of SeptiCyte RAPID was based on testing samples from retrospective (n=356) and prospective (n=63) patient cohorts. Demographic characteristics of the study cohorts are presented in **Table 1**. The majority of patients were recruited over the dates Jan 2011 - Dec 2013 (MARS), May 2014 - Apr 2015 (VENUS) and Mar - Aug 2016 (VENUS Supplement), which largely predate the Sepsis-3 definition [28]. Sepsis, SIRS or indeterminate diagnoses were assigned by RPD under the Sepsis-2 definition, although reanalysis under Sepsis-3 was also performed and is described below. When compared to patients with SIRS, patients adjudicated as septic tended to be older (p = 0.016). There were no significant differences by sex or race/ethnicity. Although the patients were evaluated in intensive care, they originated from a variety of hospital locations. Patients coming from hospital wards exhibited a higher proportion of sepsis relative to patients coming from the ED (p = 0.003), the post-anesthesia unit / post-operating room (p < 0.00001), or the ICU / cardiac care unit (p = 0.02).

**Table 1.**
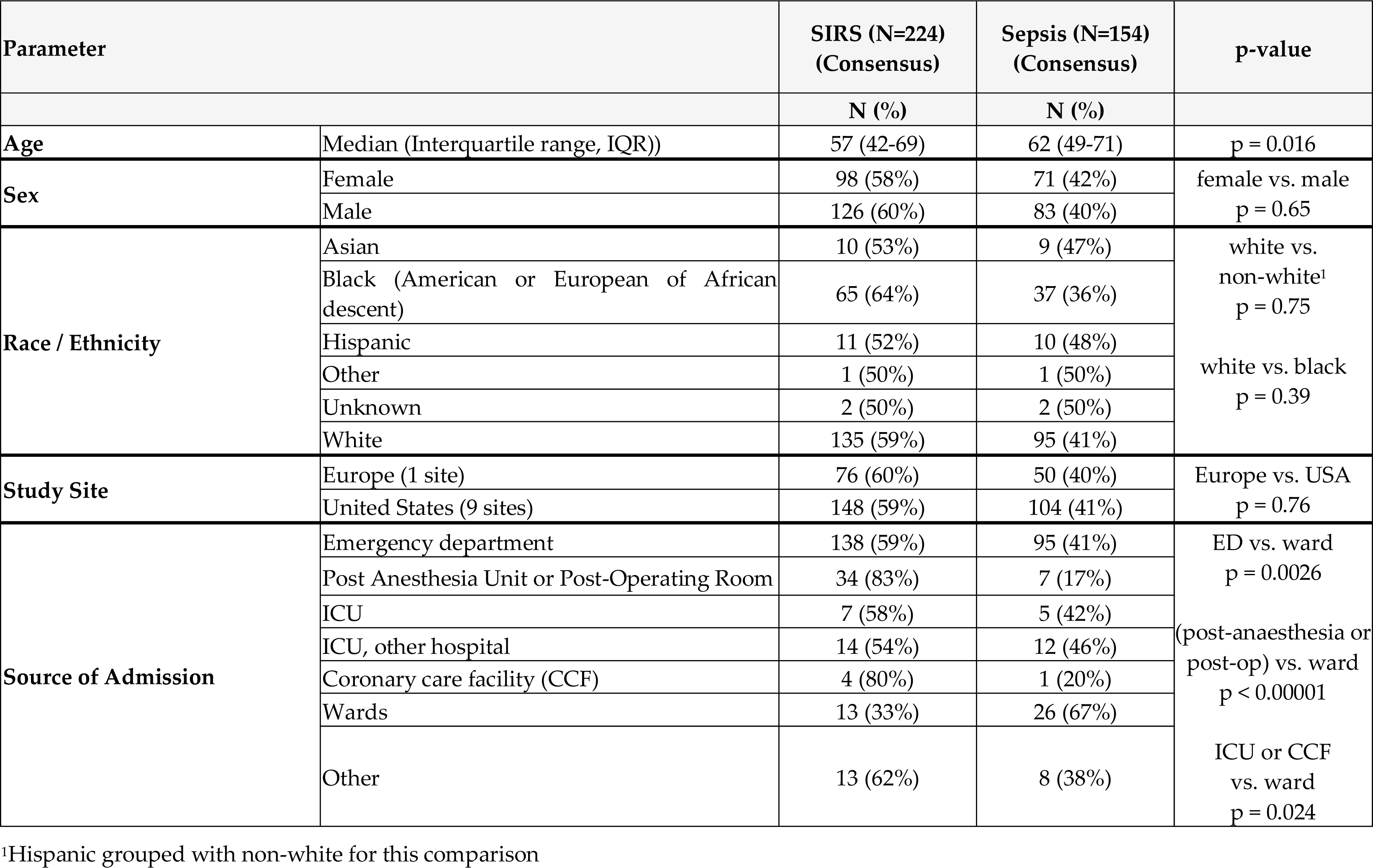
Demographic characteristics of study population. There were 41 (9.8%) indeterminates by consensus RPD, which are not shown in this table or included in the p-value calculations. Student’s t-test was used to calculate p-values for the indicated comparisons.

| Parameter |  | SIRS (N=224)<br>(Consensus) | Sepsis (N=154)<br>(Consensus) | p-value |
| --- | --- | --- | --- | --- |
|  |  | N (%) | N (%) |  |
| Age | Median (Interquartile range, IQR)) | 57 (42-69) | 62 (49-71) | p = 0.016 |
| Sex | Female | 98 (58%) | 71 (42%) | female vs. male<br>p = 0.65 |
|  | Male | 126 (60%) | 83 (40%) |  |
| Race / Ethnicity | Asian | 10 (53%) | 9 (47%) | white vs.<br>non-white<sup>1</sup><br>p = 0.75 |
|  | Black (American or European of African descent) | 65 (64%) | 37 (36%) |  |
|  | Hispanic | 11 (52%) | 10 (48%) |  |
|  | Other | 1 (50%) | 1 (50%) | white vs. black<br>p = 0.39 |
|  | Unknown | 2 (50%) | 2 (50%) |  |
|  | White | 135 (59%) | 95 (41%) |  |
| Study Site | Europe (1 site) | 76 (60%) | 50 (40%) | Europe vs. USA<br>p = 0.76 |
|  | United States (9 sites) | 148 (59%) | 104 (41%) |  |
| Source of Admission | Emergency department | 138 (59%) | 95 (41%) | ED vs. ward<br>p = 0.0026 |
|  | Post Anesthesia Unit or Post-Operating Room | 34 (83%) | 7 (17%) |  |
|  | ICU | 7 (58%) | 5 (42%) |  |
|  | ICU, other hospital | 14 (54%) | 12 (46%) | (post-anaesthesia or<br>post-op) vs. ward<br>p < 0.00001 |
|  | Coronary care facility (CCF) | 4 (80%) | 1 (20%) |  |
|  | Wards | 13 (33%) | 26 (67%) |  |
|  | Other | 13 (62%) | 8 (38%) | ICU or CCF<br>vs. ward<br>p = 0.024 |

<sup>1</sup>Hispanic grouped with non-white for this comparison

Clinical characteristics of the study cohorts are presented in **Tables 2 and 3**. Of particular note are positive culture results of high statistical significance. Positive results for blood+, urine+ and blood+/urine+ cultures were very highly associated with RPD diagnoses of sepsis as opposed to SIRS, with p-values of <0.00001, <0.00001, and <0.001 respectively. Also, positive results for blood+, urine+, and blood+/urine+ cultures were associated with elevated SeptiScores, especially SeptiScores in Band 4 (see Methods for Band definitions). Of the 48 patients that were called septic by RPD with positive blood+ or blood+/urine+ culture results, 34/48 (71%) of SeptiScores fell in band 4 and 9/48 (19%) fell in band 3. Of the 42 patients with positive blood+ culture results, only one had a SeptiScore in Band 1; this patient was diagnosed as SIRS by RPD, with the blood culture result being considered a probable contaminant by the attending physician and the RPD panelists.

**Table 2.**
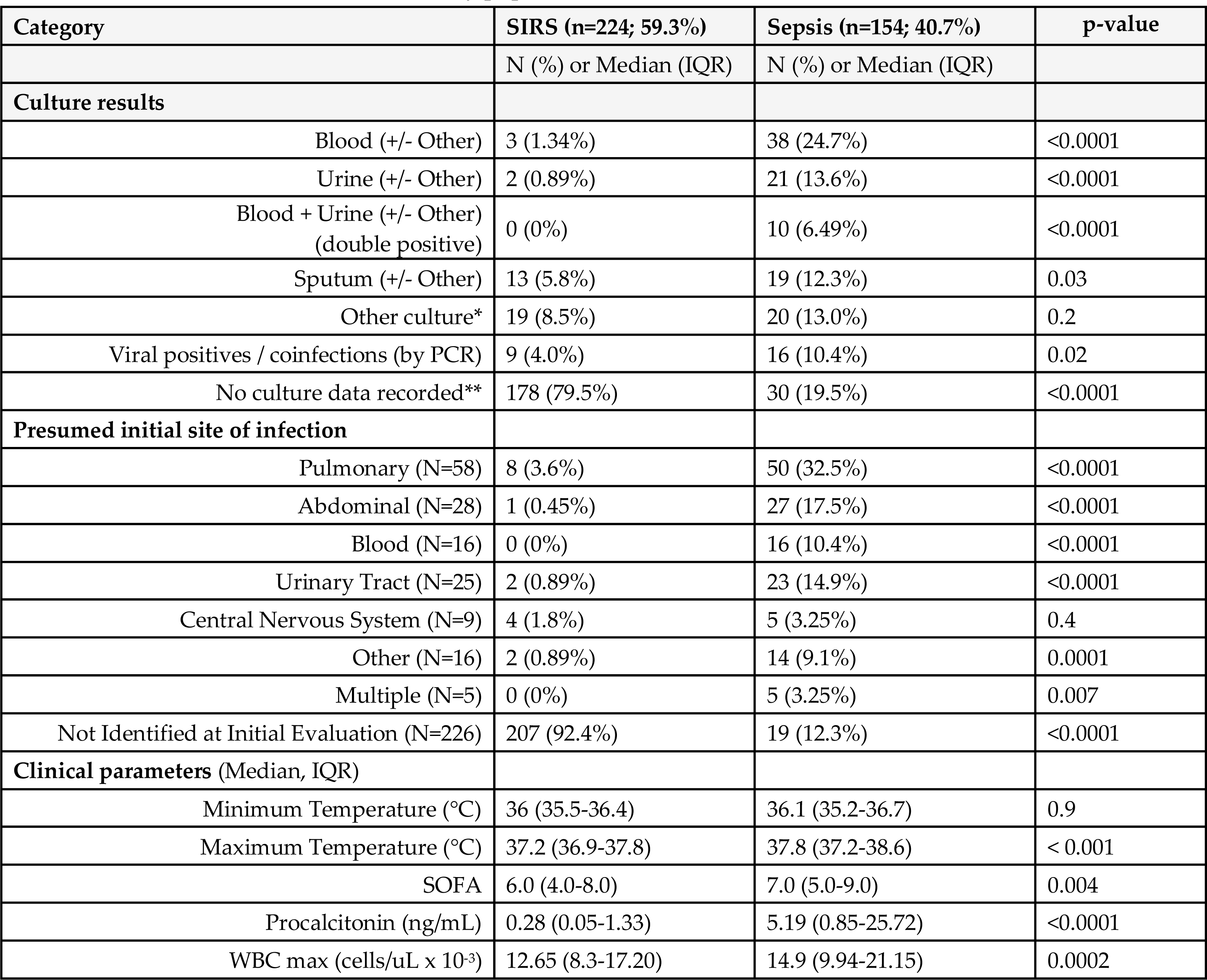
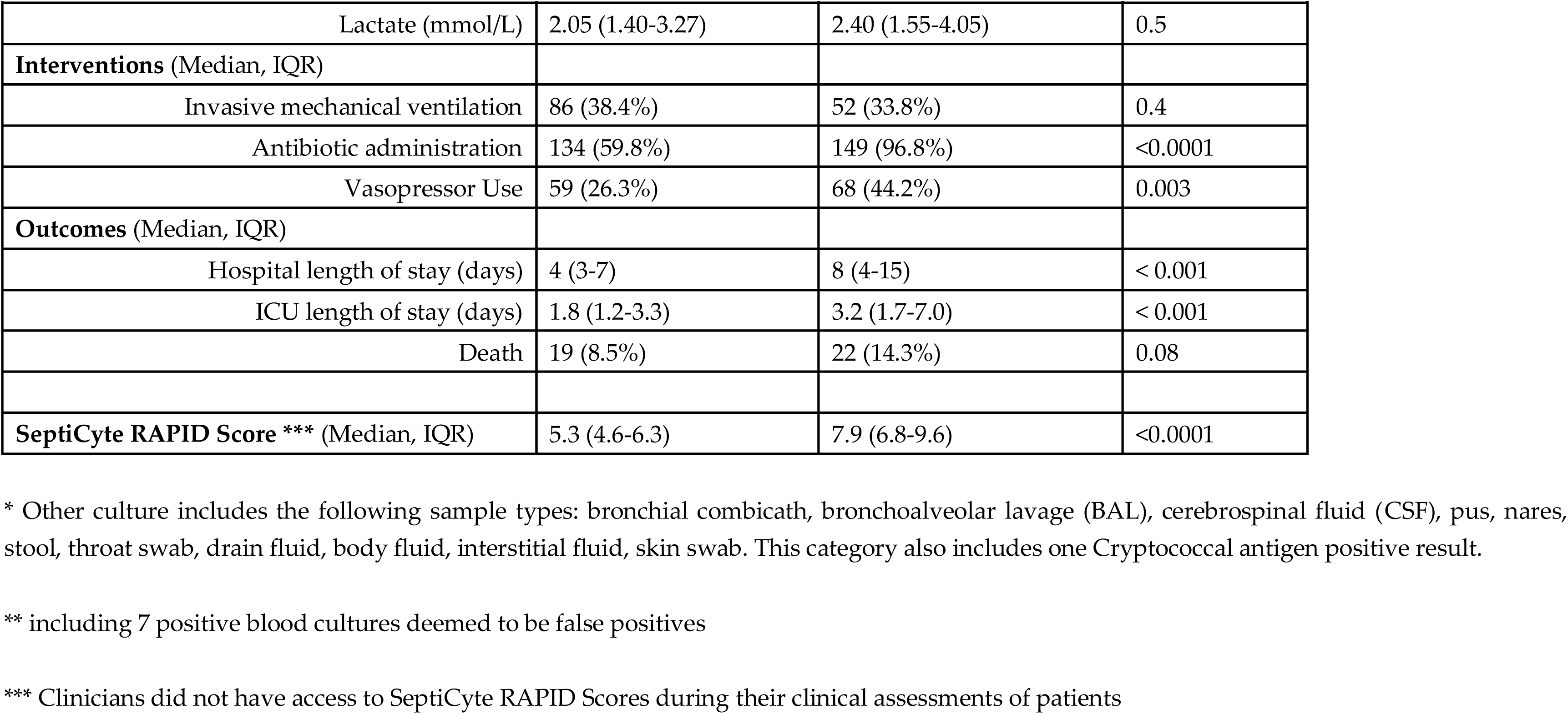
Clinical characteristics of the study population. Consensus RPD was used as the comparator. There were 41/419 (9.8%) indeterminates by consensus RPD, which are not shown in this table or included in the p-value calculations. For categorical variables, the chi-squared test was used to calculate p-values for the difference in prevalence (%) of observations of each type across the SIRS and sepsis categories. For continuous variables, Welch’s two sample t-test was used to calculate p-values across the SIRS and sepsis categories. Abbreviation: n.a., not appropriate.

**Table 3.**
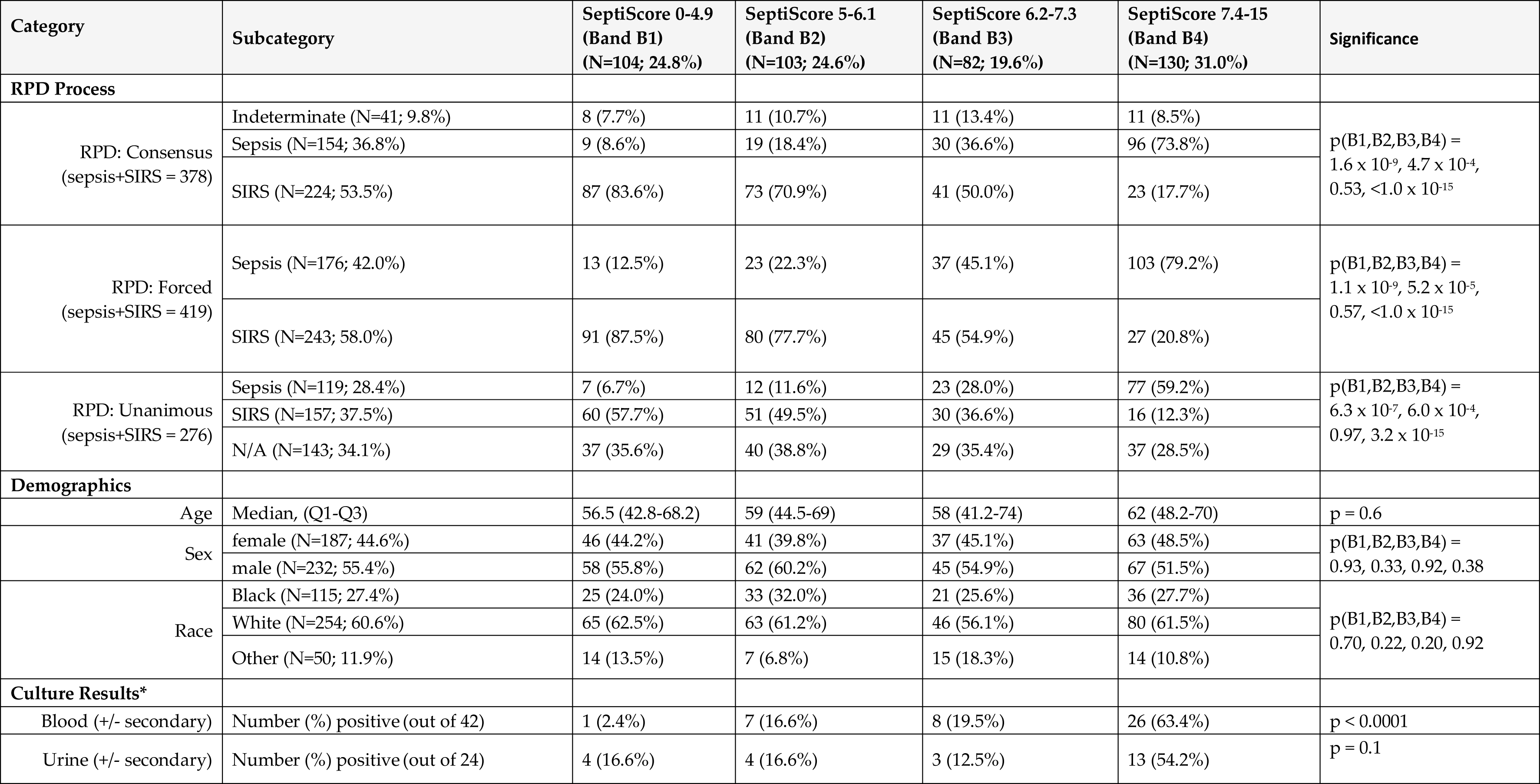
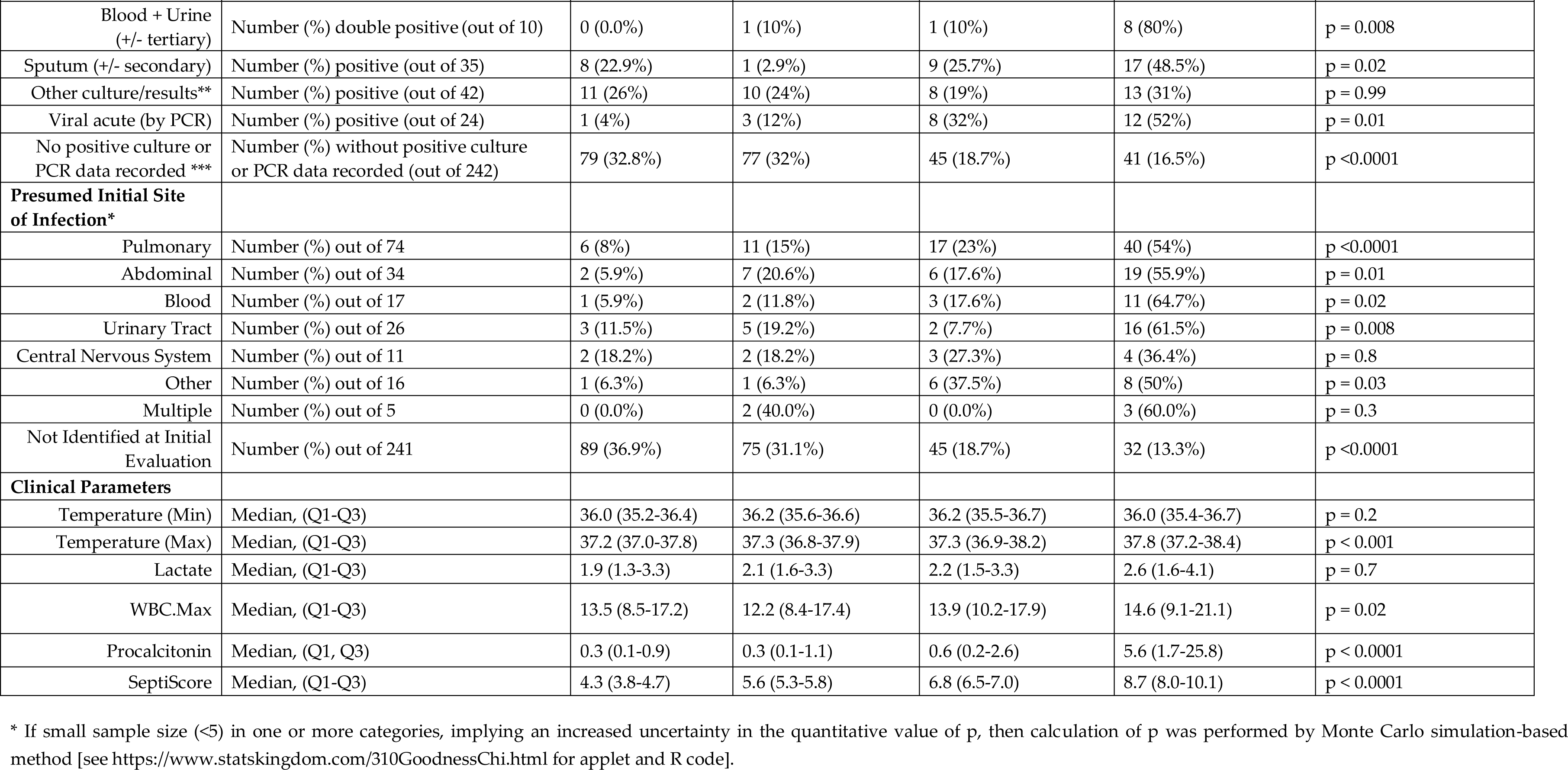
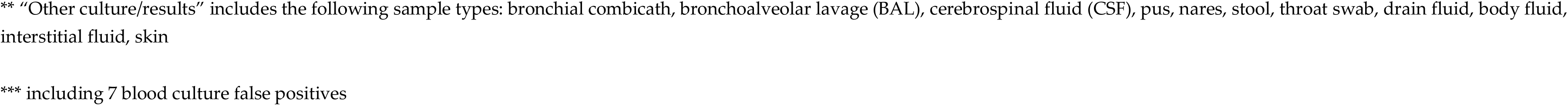
Characteristics of all patients stratified by RPD, demographic and clinical parameters, and SeptiScore band. A chi-squared test or ANOVA was used, for categorical variables or continuous variables, respectively, to estimate p-values across or within the SeptiScore bands as appropriate. In the chi-squared analyses, p(Bi, i=1…4) means the probability of the observed vs. expected distributions of patients across sub-categories (e.g. male, female; or Black, White, Other) within band Bi.

| Category | Subcategory | SeptiScore 0-4.9<br>(Band B1)<br>(N=104; 24.8%) | SeptiScore 5-6.1<br>(Band B2)<br>(N=103; 24.6%) | SeptiScore 6.2-7.3<br>(Band B3)<br>(N=82; 19.6%) | SeptiScore 7.4-15<br>(Band B4)<br>(N=130; 31.0%) | Significance |
| --- | --- | --- | --- | --- | --- | --- |
| RPD Process |  |  |  |  |  |  |
| RPD: Consensus<br>(sepsis+SIRS = 378) | Indeterminate (N=41; 9.8%) | 8 (7.7%) | 11 (10.7%) | 11 (13.4%) | 11 (8.5%) | p(B1,B2,B3,B4) =<br>1.6 x 10<sup>-9</sup>, 4.7 x 10<sup>-4</sup>,<br>0.53, <1.0 x 10<sup>-15</sup> |
|  | Sepsis (N=154; 36.8%) | 9 (8.6%) | 19 (18.4%) | 30 (36.6%) | 96 (73.8%) |  |
|  | SIRS (N=224; 53.5%) | 87 (83.6%) | 73 (70.9%) | 41 (50.0%) | 23 (17.7%) |  |
| RPD: Forced<br>(sepsis+SIRS = 419) | Sepsis (N=176; 42.0%) | 13 (12.5%) | 23 (22.3%) | 37 (45.1%) | 103 (79.2%) | p(B1,B2,B3,B4) =<br>1.1 x 10<sup>-9</sup>, 5.2 x 10<sup>-5</sup>,<br>0.57, <1.0 x 10<sup>-15</sup> |
|  | SIRS (N=243; 58.0%) | 91 (87.5%) | 80 (77.7%) | 45 (54.9%) | 27 (20.8%) |  |
| RPD: Unanimous<br>(sepsis+SIRS = 276) | Sepsis (N=119; 28.4%) | 7 (6.7%) | 12 (11.6%) | 23 (28.0%) | 77 (59.2%) | p(B1,B2,B3,B4) =<br>6.3 x 10<sup>-7</sup>, 6.0 x 10<sup>-4</sup>,<br>0.97, 3.2 x 10<sup>-15</sup> |
|  | SIRS (N=157; 37.5%) | 60 (57.7%) | 51 (49.5%) | 30 (36.6%) | 16 (12.3%) |  |
|  | N/A (N=143; 34.1%) | 37 (35.6%) | 40 (38.8%) | 29 (35.4%) | 37 (28.5%) |  |
| Demographics |  |  |  |  |  |  |
| Age | Median, (Q1-Q3) | 56.5 (42.8-68.2) | 59 (44.5-69) | 58 (41.2-74) | 62 (48.2-70) | p = 0.6 |
| Sex | female (N=187; 44.6%) | 46 (44.2%) | 41 (39.8%) | 37 (45.1%) | 63 (48.5%) | p(B1,B2,B3,B4) =<br>0.93, 0.33, 0.92, 0.38 |
|  | male (N=232; 55.4%) | 58 (55.8%) | 62 (60.2%) | 45 (54.9%) | 67 (51.5%) |  |
| Race | Black (N=115; 27.4%) | 25 (24.0%) | 33 (32.0%) | 21 (25.6%) | 36 (27.7%) | p(B1,B2,B3,B4) =<br>0.70, 0.22, 0.20, 0.92 |
|  | White (N=254; 60.6%) | 65 (62.5%) | 63 (61.2%) | 46 (56.1%) | 80 (61.5%) |  |
|  | Other (N=50; 11.9%) | 14 (13.5%) | 7 (6.8%) | 15 (18.3%) | 14 (10.8%) |  |
| Culture Results* |  |  |  |  |  |  |
| Blood (+/- secondary) | Number (%) positive (out of 42) | 1 (2.4%) | 7 (16.6%) | 8 (19.5%) | 26 (63.4%) | p < 0.0001 |
| Urine (+/- secondary) | Number (%) positive (out of 24) | 4 (16.6%) | 4 (16.6%) | 3 (12.5%) | 13 (54.2%) | p = 0.1 |
| Blood + Urine<br>(+/- tertiary) | Number (%) double positive (out of 10) | 0 (0.0%) | 1 (10%) | 1 (10%) | 8 (80%) | p = 0.008 |
| Sputum (+/- secondary) | Number (%) positive (out of 35) | 8 (22.9%) | 1 (2.9%) | 9 (25.7%) | 17 (48.5%) | p = 0.02 |
| Other culture/results** | Number (%) positive (out of 42) | 11 (26%) | 10 (24%) | 8 (19%) | 13 (31%) | p = 0.99 |
| Viral acute (by PCR) | Number (%) positive (out of 24) | 1 (4%) | 3 (12%) | 8 (32%) | 12 (52%) | p = 0.01 |
| No positive culture or<br>PCR data recorded *** | Number (%) without positive culture<br>or PCR data recorded (out of 242) | 79 (32.8%) | 77 (32%) | 45 (18.7%) | 41 (16.5%) | p <0.0001 |
| **Presumed Initial Site**<br>**of Infection\*** |  |  |  |  |  |  |
| Pulmonary | Number (%) out of 74 | 6 (8%) | 11 (15%) | 17 (23%) | 40 (54%) | p <0.0001 |
| Abdominal | Number (%) out of 34 | 2 (5.9%) | 7 (20.6%) | 6 (17.6%) | 19 (55.9%) | p = 0.01 |
| Blood | Number (%) out of 17 | 1 (5.9%) | 2 (11.8%) | 3 (17.6%) | 11 (64.7%) | p = 0.02 |
| Urinary Tract | Number (%) out of 26 | 3 (11.5%) | 5 (19.2%) | 2 (7.7%) | 16 (61.5%) | p = 0.008 |
| Central Nervous System | Number (%) out of 11 | 2 (18.2%) | 2 (18.2%) | 3 (27.3%) | 4 (36.4%) | p = 0.8 |
| Other | Number (%) out of 16 | 1 (6.3%) | 1 (6.3%) | 6 (37.5%) | 8 (50%) | p = 0.03 |
| Multiple | Number (%) out of 5 | 0 (0.0%) | 2 (40.0%) | 0 (0.0%) | 3 (60.0%) | p = 0.3 |
| Not Identified at Initial<br>Evaluation | Number (%) out of 241 | 89 (36.9%) | 75 (31.1%) | 45 (18.7%) | 32 (13.3%) | p <0.0001 |
| **Clinical Parameters** |  |  |  |  |  |  |
| Temperature (Min) | Median, (Q1-Q3) | 36.0 (35.2-36.4) | 36.2 (35.6-36.6) | 36.2 (35.5-36.7) | 36.0 (35.4-36.7) | p = 0.2 |
| Temperature (Max) | Median, (Q1-Q3) | 37.2 (37.0-37.8) | 37.3 (36.8-37.9) | 37.3 (36.9-38.2) | 37.8 (37.2-38.4) | p < 0.001 |
| Lactate | Median, (Q1-Q3) | 1.9 (1.3-3.3) | 2.1 (1.6-3.3) | 2.2 (1.5-3.3) | 2.6 (1.6-4.1) | p = 0.7 |
| WBC.Max | Median, (Q1-Q3) | 13.5 (8.5-17.2) | 12.2 (8.4-17.4) | 13.9 (10.2-17.9) | 14.6 (9.1-21.1) | p = 0.02 |
| Procalcitonin | Median, (Q1, Q3) | 0.3 (0.1-0.9) | 0.3 (0.1-1.1) | 0.6 (0.2-2.6) | 5.6 (1.7-25.8) | p < 0.0001 |
| SeptiScore | Median, (Q1-Q3) | 4.3 (3.8-4.7) | 5.6 (5.3-5.8) | 6.8 (6.5-7.0) | 8.7 (8.0-10.1) | p < 0.0001 |

\* If small sample size (<5) in one or more categories, implying an increased uncertainty in the quantitative value of p, then calculation of p was performed by Monte Carlo simulation-based method [see https://www.statskingdom.com/310GoodnessChi.html for applet and R code].
\*\* “Other culture/results” includes the following sample types: bronchial combicath, bronchoalveolar lavage (BAL), cerebrospinal fluid (CSF), pus, nares, stool, throat swab, drain fluid, body fluid, interstitial fluid, skin
\*\*\* including 7 blood culture false positives

A taxonomic analysis of the culture-confirmed pathogens for the sepsis cases is presented in the Supplementary Information, Sections 8-9. In our study cohorts, SeptiScore does not appear to be significantly affected by the type of pathogen (bacterial, viral, fungal) underlying a sepsis event. In particular, we observed no significant difference (p = 0.264) between the SeptiScore distributions for septic patients with Gram-positive infections (n=55) versus Gram-negative infections (n=36).

### 3.3 SeptiCyte RAPID performance under Sepsis-2

#### 3.3.1 ROC curve analyses

Receiver operating characteristic (ROC) curve analyses for sepsis vs SIRS were performed for the retrospective cohort (**Figure 1A**), the prospective cohort (**Figure 1B**), and the combined retrospective + prospective cohorts (**Figure 1C**). Performance estimates for the retrospective cohort ranged from AUC 0.82-0.85 (depending upon RPD method) and were statistically indistinguishable both from each other, and from the AUC values previously obtained for SeptiCyte LAB [23]. Performance estimates for the prospective cohort ranged from AUC 0.86-0.95 (depending upon RPD method) and again were statistically indistinguishable from each other.

DeLong’s test [33] showed no significant difference between the ROC curves from Figure 1A (retrospective cohort) vs. 1B (prospective cohort), when either forced or consensus RPD was used as comparator. When unanimous RPD was used as comparator, DeLong’s test gave p = 0.013. For the retrospective + prospective dataset, using consensus RPD as comparator and assuming a binary distinction between sepsis and SIRS, the relationship between cutpoint, sensitivity, and specificity is shown in **Figure 1D**.

#### 3.3.2 Sepsis probability distributions

The distribution of SeptiScores for septic patients falls above that for patients with SIRS (**Figure 2A**). A sliding window analysis (**Figure 2B**) was used to calculate the probability of sepsis as a function of SeptiScore (**Figure 2C**). Patients with higher SeptiScores had higher probabilities of sepsis. As described in Methods, the 0-15 SeptiScore range has been divided into four probability bands (**Figure 2D**) defined by the pre-set band boundaries of 4.95, 6.15 and 7.45. **Figure 3** and **Table 4** present SeptiScores for the complete (retrospective + prospective) cohort, and for the retrospective and prospective cohorts separately, after parsing into the four bands. Using consensus RPD as comparator, Band 1 had negative predictive value 0.91 (sensitivity 0.94), and Band 4 had positive predictive value 0.81 (specificity 0.90). Chi-squared analyses (**Table 4**) indicated that, after sorting patients into the four SeptiScore bands, no significant influences were found with respect to the baseline demographic criteria or sex, age, race or ethnicity (**Table 3**).

**Figure 2.**
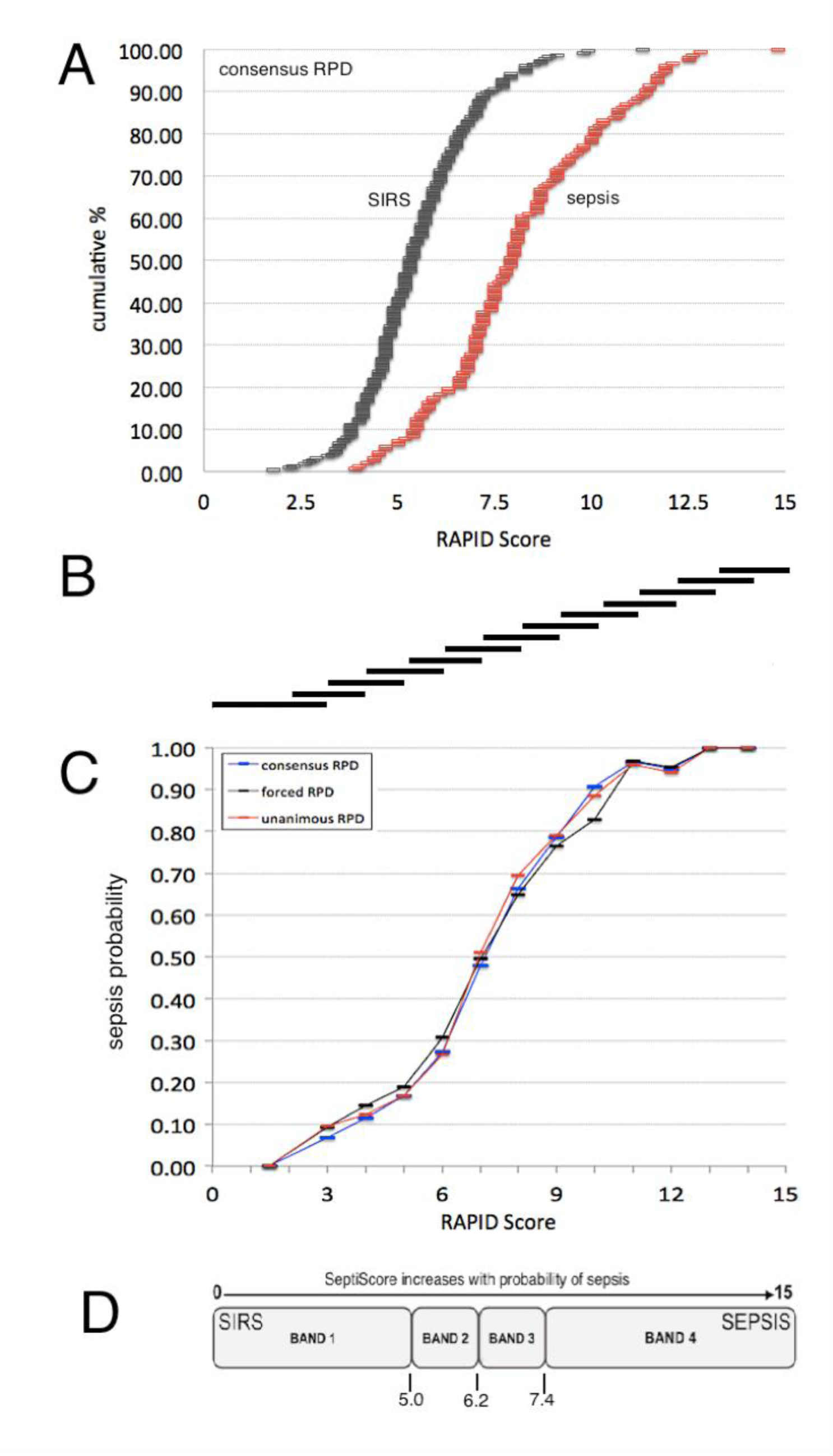
Probability of sepsis as a function of SeptiScore. **(A)** Cumulative distributions of the SeptiScore for SIRS patients (black) and sepsis patients (red), as evaluated by consensus RPD. **(B)** Basis of sliding window calculation of P(sepsis) across the 0-15 SeptiScore range. A sliding window was defined to be 3 score-units wide initially (from 0 to 3), and 2-score-units wide thereafter. It was shifted in 1-unit increments. The number of sepsis and SIRS in each placement of the window was counted, and P(sepsis) calculated as N_sepsis_ / (N_sepsis_ + N_SIRS_). **(C)** Probability of sepsis as function of SeptiScore, from the sliding window analysis. Key: black curve, forced RPD; blue curve, consensus RPD; red curve, unanimous RPD. **(D)** Division of SeptiScore range into four sepsis probability bands.

**Figure 3.**
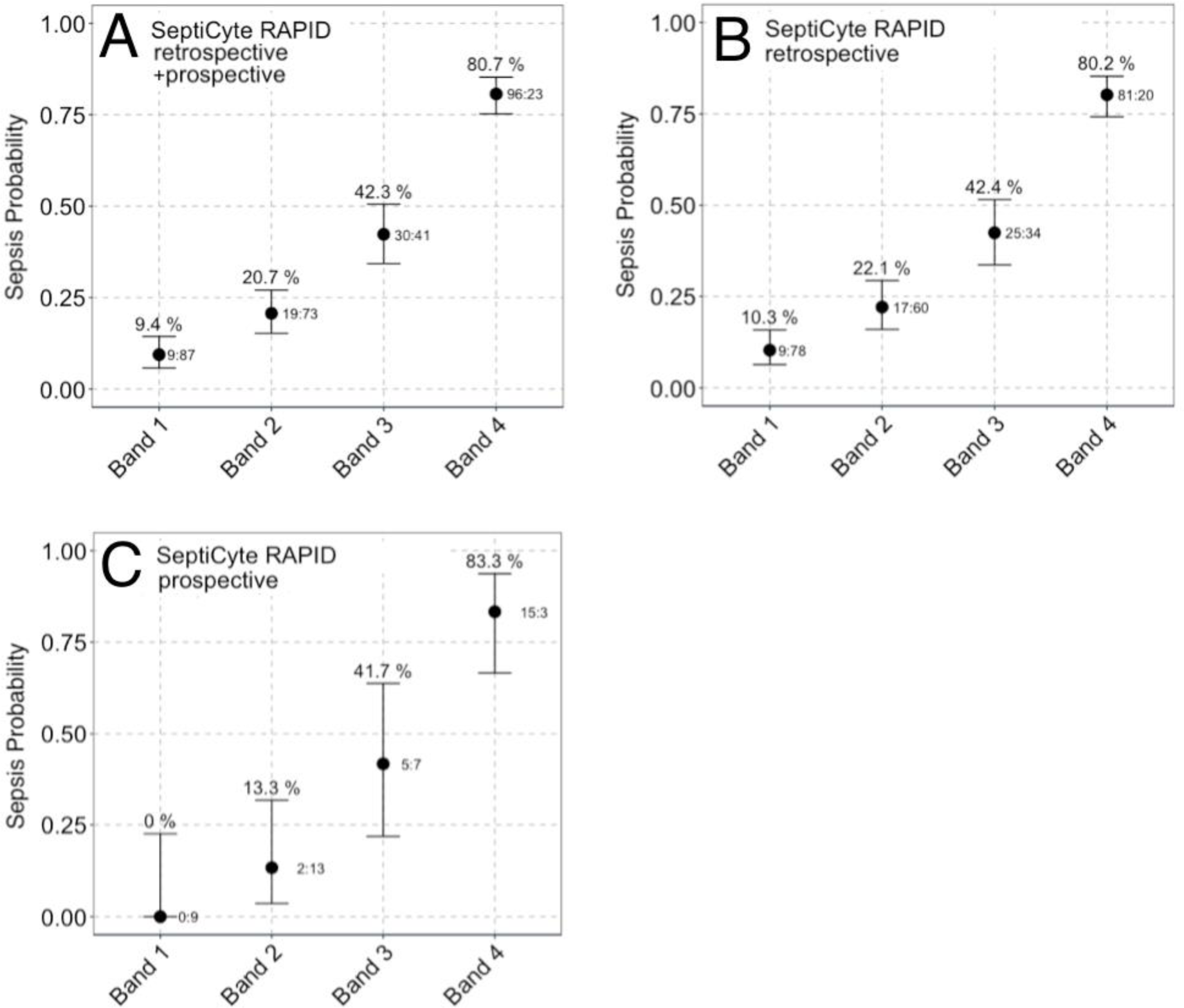
Comparison of sepsis probabilities per band for **(A)** SeptiCyte RAPID combined retrospective + prospective cohorts (N=378), **(B)** SeptiCyte RAPID retrospective cohort (N=324), and **(C)** SeptiCyte RAPID prospective cohort (N=54). Assignment of sepsis or SIRS was by consensus RPD. Subjects called “indeterminate” by consensus RPD are not included. The ratio to the right of each whisker (x:y) indicates the number of patients called as sepsis (x) or SIRS (y) in the associated SeptiScore band. The number above each whisker describes the percentage of sepsis calls in each band. The error bars represent the lower and upper bounds of the 80% confidence interval for the sample proportion, calculated using the exact binomial formula.

**Table 4.** Interpretation of SeptiCyte RAPID scores. This table provides the performance metrics for SeptiScores falling within each band. The consensus RPD method was used as comparator in all analyses.

| SeptiScore Band | Cohort | Average Probability |  | Sepsis Likelihood Ratio | Percentage of Cohort |
| --- | --- | --- | --- | --- | --- |
|  |  | SIRS | Sepsis |  |  |
| Band 4<br>(7.4 – 15)<br>High Risk of Sepsis | Retrospective | 20 (20%) | 81 (80%) | 5.89 | 31% |
|  | Prospective | 3 (17%) | 15 (83%) | 7.27 | 33% |
|  | Combined | 23 (19%) | 96 (81%) | 6.07 | 32% |
| Band 3<br>(6.2 – 7.3) | Retrospective | 34 (58%) | 25 (42%) | 1.07 | 18% |
|  | Prospective | 7 (58%) | 5 (42%) | 1.04 | 22% |
|  | Combined | 41 (58%) | 30 (42%) | 1.06 | 19% |
| Band 2<br>(5.0 – 6.1) | Retrospective | 60 (78%) | 17 (22%) | 0.41 | 24% |
|  | Prospective | 13 (87%) | 2 (13%) | 0.22 | 28% |
|  | Combined | 73 (79%) | 19 (21%) | 0.38 | 24% |
| Band 1<br>(0 – 4.9)<br>Low Risk of Sepsis | Retrospective | 78 (90%) | 9 (10%) | 0.17 | 27% |
|  | Prospective | 9 (100%) | 0 (0%) | 0.00 | 17% |
|  | Combined | 87 (91%) | 9 (9%) | 0.15 | 25% |

#### 3.3.3 Multivariable analysis

Under the Sepsis-2 framework, we asked if SeptiCyte RAPID could provide diagnostic clinical utility for discriminating between sepsis and SIRS patients, beyond that provided by other clinical variables and laboratory assessments available on the first day of ICU admission. Fourteen variables in addition to SeptiCyte RAPID were examined (see Supplementary Information, Sections 4-5). We evaluated all 32,767 possible combinations of the fifteen variables, and performance (sepsis vs. SIRS under the Sepsis-2 framework) was assessed by AUC against consensus RPD. Procalcitonin was included in this analysis, as well as lactate which is commonly used in determining which patients should receive sepsis treatment bundles [35, 36]. **Figure 4A** shows the results of the multivariable analysis for the combined retrospective and prospective cohorts, while **Figure 4B** shows the comparable results for just the prospective cohort. The AUC distributions for the prospective cohort appear somewhat high compared to those for the full patient cohort, possibly because of overfitting (the prospective cohort has only 54 patients with SIRS or sepsis by consensus RPD, as compared to 154 in the full patient cohort). With respect to individual variables, SeptiScore was found to rank highest by AUC. When combinations of clinical variables were considered, those containing SeptiScore were found to have higher AUCs than all combinations lacking SeptiScore.

**Figure 4.**
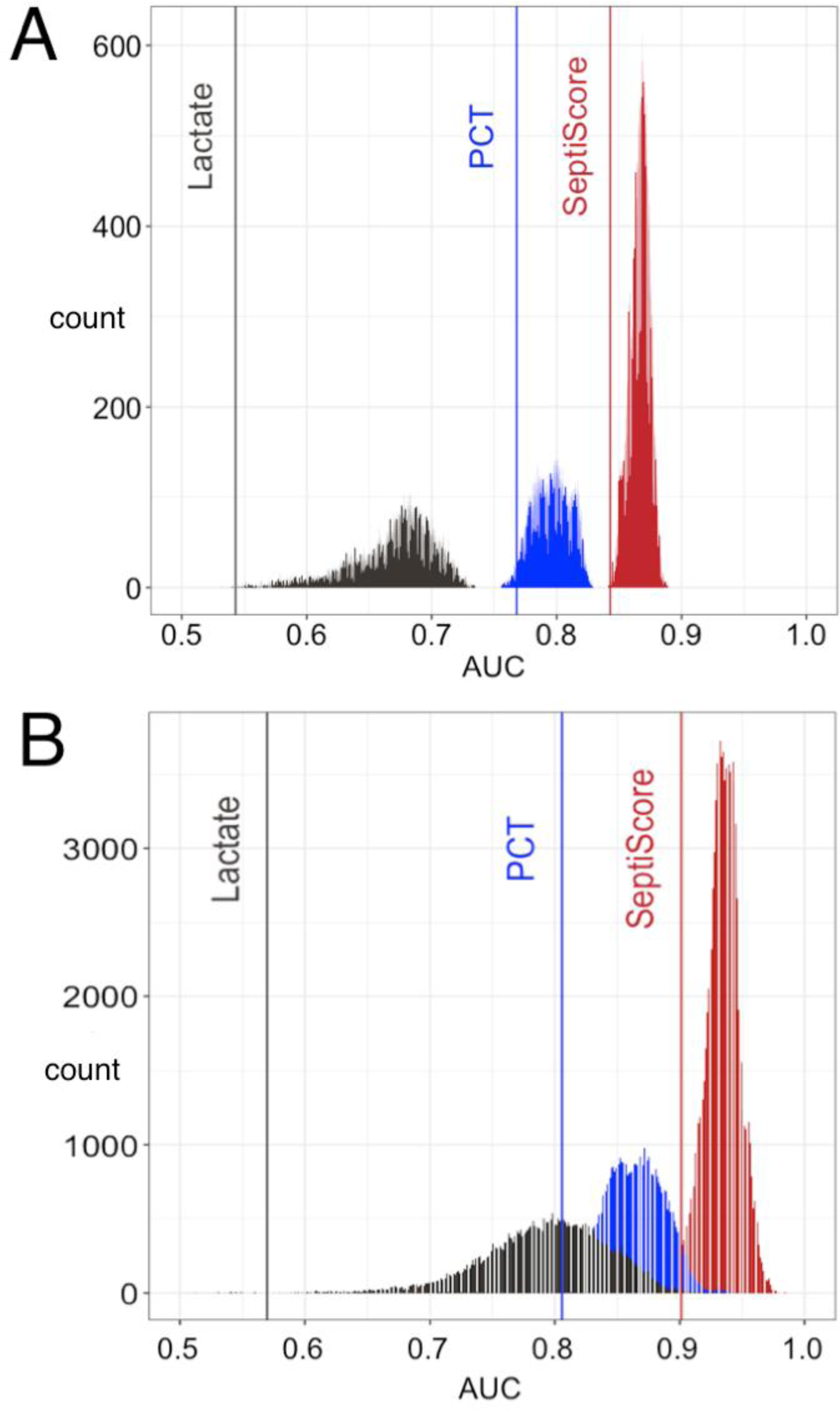
Comparison of lactate, procalcitonin and SeptiScore, without or with additional clinical variables, for discrimination of sepsis vs. SIRS. **(A)** Data from the combined retrospective and prospective cohorts. **(B)** Data from just the prospective cohort. Area under curve (AUC) distributions are shown for all 32,767 possible logistic combinations of the following variables: age, race (African-American or not), sex, MAP max, T min, T max, HR min, HR max, WBC min, WBC max, glucose max, lactate, procalcitonin, SeptiScore, num.SIRS. (Abbreviations: MAP, mean arterial pressure; T, core body temperature; HR, heart rate; WBC, white blood cell count; num.SIRS, number of SIRS criteria met.) Performance was assessed against consensus RPD. Key: grey vertical line, lactate alone; grey distribution, lactate combined with other clinical variables except SeptiScore or PCT; blue vertical line, PCT alone; blue distribution, PCT combined with other clinical variables except SeptiScore; red vertical line, SeptiScore alone; red distribution, SeptiScore combined with other clinical variables.

### 3.4 SeptiCyte RAPID performance under Sepsis-3

In 2016, Singer et al. [28] proposed that the Sepsis-2 definition be replaced with Sepsis-3 which identifies sepsis as “life-threatening organ dysfunction caused by a dysregulated host response to infection”. We compared the performance of SeptiCyte RAPID for discriminating sepsis vs. non-sepsis under the Sepsis-2 vs. Sepsis-3 definitions. **Figure 5** displays 2×2 tables for classifying patients in terms of (+/− organ dysfunction) x (+/− infection). Panels A, B employ the Sepsis-2 definition, while Panels C, D employ the Sepsis-3 definition. In each panel, the numbers in red indicate septic patients, according to whether organ dysfunction was indicated by complete SOFA score ≥2 (Panels A, C) or by partial SOFA score ≥2 (Panels B, D). A striking feature of these tables is that they reveal nearly all the patients in the study cohort display some amount of organ dysfunction, indicated by SOFA ≥2 (256/267 = 95.9% of patients) or pSOFA ≥2 (316/372 = 84.9% of patients). This however might be expected, since all patients in the cohort were admitted to ICU.

**Figure 5.**
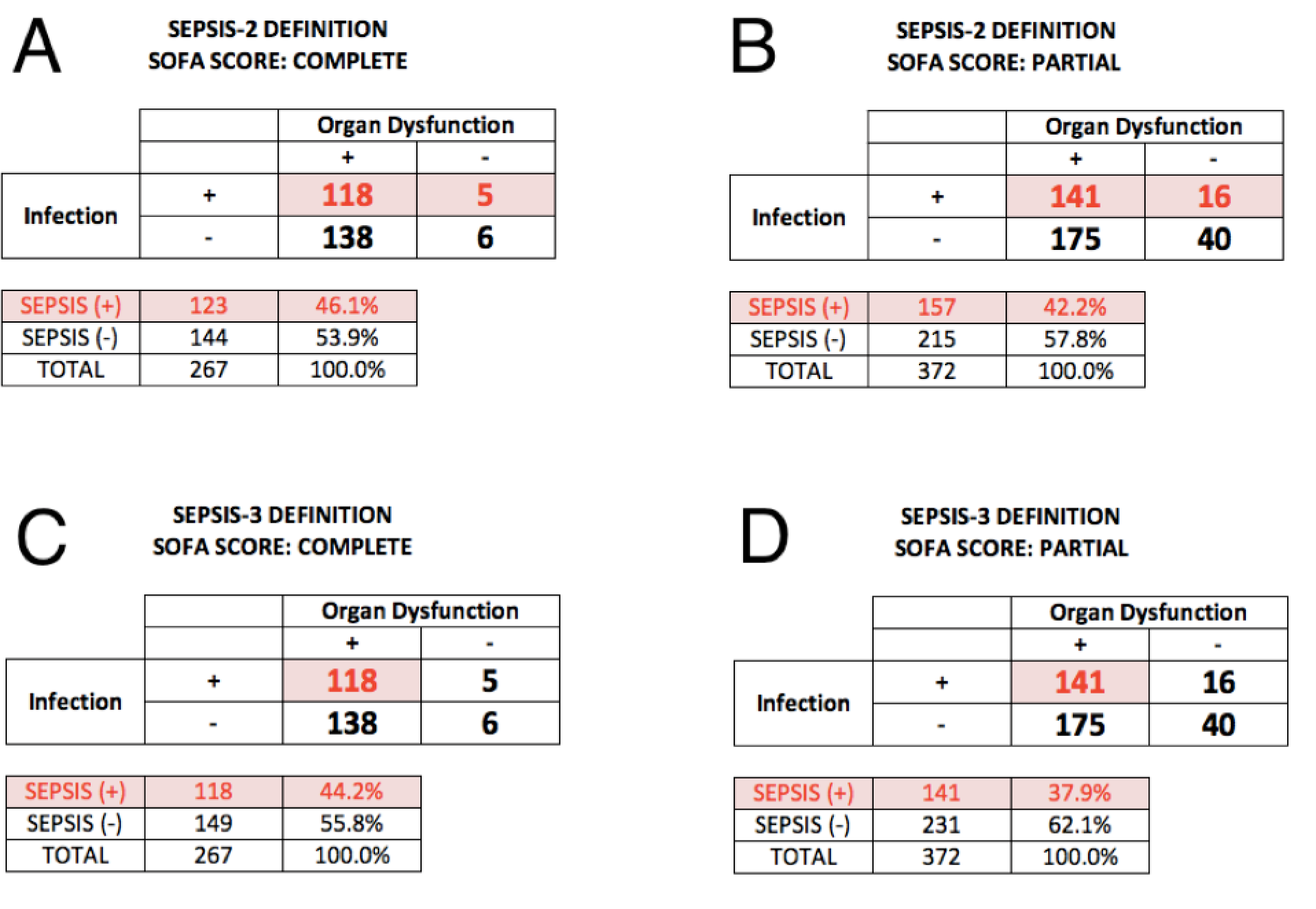
Classification of patients in terms of 2×2 tables (+/− organ dysfunction) x (+/− infection), according to the Sepsis-2 vs. Sepsis-3 definitions. The numbers in red indicate the septic patients according to the respective definitions. **(A)** Sepsis-2 definition, with complete SOFA score ≥2 indicating organ dysfunction. **(B)** Sepsis-2 definition, with partial SOFA score ≥2 indicating organ dysfunction. **(C)** Sepsis-3 definition, with complete SOFA score ≥2 indicating organ dysfunction. **(D)** Sepsis-3 definition, with partial SOFA score ≥2 indicating organ dysfunction.

**Figure 6** builds upon Figure 5, using ROC analysis to evaluate SeptiCyte RAPID performance for each patient grouping. Performance under the Sepsis-2 vs. Sepsis-3 definitions is compared when organ dysfunction is indicated either by complete SOFA score ≥2 (panel **A**) or by partial SOFA score ≥2 (panel **B**). The ROC curves were found statistically equivalent across all these comparisons, with AUC values ranging from 0.807 to 0.820. Additional analyses, concerning the relative independence of SeptiScore on the extent of organ dysfunction, are presented in Supplementary Material, Section 10.

**Figure 6.**
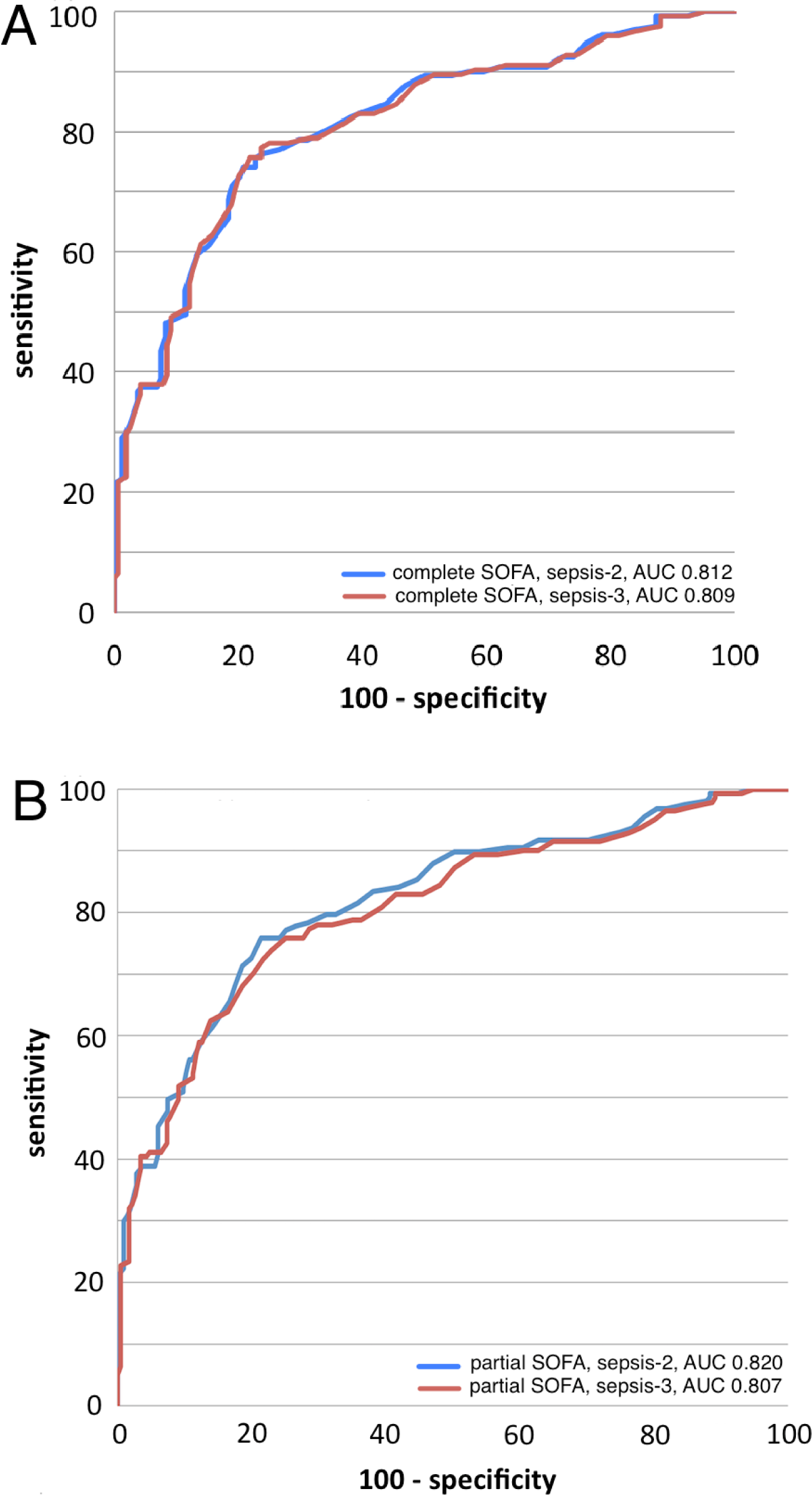
SeptiCyte RAPID performance by ROC analysis. **(A)** ROC curves for discriminating sepsis vs. non-sepsis under the Sepsis-2 vs. Sepsis-3 definitions when organ dysfunction is indicated by complete SOFA score ≥2. Blue curve: sepsis-2 definition, AUC 0.812. Red curve: sepsis-3 definition, AUC 0.809. **(B)** ROC curves for discriminating sepsis vs. non-sepsis under the Sepsis-2 vs. Sepsis-3 definitions when organ dysfunction is indicated by partial SOFA score ≥2. Blue curve: sepsis-2 definition, AUC 0.820. Red curve: sepsis-3 definition, AUC 0.807.

## 4. Discussion

We have presented data validating the use of SeptiCyte RAPID for discriminating sepsis from SIRS and for estimating the probability of sepsis in critically ill adult patients, within a clinically actionable time frame. Analysis was performed under both the Sepsis-2 and Sepsis-3 frameworks. Patients in this study came from the ED, post-anesthesia unit, post-operating rooms and wards, and were tested in the ICU. For the full (retrospective + prospective) cohort, diagnostic performance of SeptiCyte RAPID under the Sepsis-2 definition was equivalent or superior to that previously reported for SeptiCyte LAB [23].

In a multivariable analysis under the Sepsis-2 framework, we examined all possible combinations of SeptiScore and up to 14 additional clinical or laboratory variables, including lactate and PCT. We found that SeptiScore alone had greater performance than any combination of variables without SeptiScore, for differentiating sepsis vs. SIRS (Figure 4). However, the analysis also indicated it should be possible to moderately boost the performance of SeptiCyte RAPID by combining the SeptiScore with other clinical parameters.

We reanalyzed our data under the Sepsis-3 framework, using the SOFA or pSOFA score as a quantitative measure of organ dysfunction [29–31], and interpreting a RPD call of sepsis as indicating probable or definite infection. The results (Figure 6) indicated that SeptiScore has high diagnostic performance for detecting sepsis (AUC 0.807-0.820) under either the Sepsis-2 or Sepsis-3 framework. Because SeptiCyte RAPID performance appears independent of organ dysfunction, we hypothesize that the SeptiCyte RAPID expression signature may be responding to some process or condition in the septic trajectory occurring earlier than organ damage, likely related to the presence of an infection. A similar hypothesis has been stated by Lukaszewski et al. [16] in connection with an independently discovered sepsis signature.

Our reanalysis under Sepsis-3 also identified a subset of 16 patients called septic by the RPD panelists under the Sepsis-2 definition, but without appreciable organ dysfunction (Figure 5B). Under the Sepsis-3 framework, these patients would not be called septic, and therefore represent a discordance between the two sepsis definitions. We are continuing to study the clinical characteristics of these patients, to understand why they were called sepsis under Sepsis-2 but not Sepsis-3. We note that Engoren et al. [37] has identified a similar discordance: in a large retrospective analysis of over 29,000 hospitalized patients with suspected infection, 44% were called septic according to Sepsis-2, and 41% called septic according to Sepsis-3, but only 23% satisfied both sets of criteria, implying poor agreement (kappa 0.213) between Sepsis-2 and Sepsis-3 in their cohort. These investigators hypothesized that Sepsis-2 and Sepsis-3 represent different phenotypes that only partially overlap.

Our study has several limitations. Our comparator method (RPD) is imperfect. In the complete (n=419) validation dataset, three expert clinicians failed to reach either a unanimous or consensus diagnosis for 41/419 (9.8%) of patients. The use of an imperfect comparator sets an upper limit on the measurable diagnostic performance of a new test [38]. Our cohort consisted of adult patients within 24 hours of ICU admission, so generalization to other patient cohorts not been established. To avoid potential confounding effects, the study excluded subjects who received therapeutic antibiotic treatment > 24 hours before ICU admission, which could comprise a significant fraction of patients transferred from the floor to ICU. Most of the study was based on re-analysis of banked samples (356/419 = 85%), while only 63/419 (15%) were prospectively collected. We have not conducted serial sampling to measure variation in SeptiCyte RAPID scores as patients move through the ICU. We have, however, previously reported strong diagnostic performance of SeptiCyte LAB in children [39], and the high correlation between SeptiCyte LAB and SeptiCyte RAPID suggests that an equivalently strong performance in children will be found for SeptiCyte RAPID. SeptiScores falling in Bands 2 or 3 do not provide definitive conclusions (i.e. very high or low probabilities) regarding the absence or presence of sepsis. Interpretation of SeptiScores in this range would be enhanced by combining with other clinical variables to adjust the post-test sepsis probabilities (see Figure 4).

During the discovery and initial validation of the PLAC8 and PLA2G7 biomarkers [22, 23], patients with a broad range of co-morbidities were examined, including septic patients with confirmed bacterial, viral, fungal infections and malignancies, and non-septic patients with non-infectious systemic inflammation of varying etiologies. The present validation cohort also included use of a broad range of prescribed medications such as immunosuppressants, anti-neoplastic drugs, antithrombotics, corticosteroids and statins. To our knowledge, SeptiCyte RAPID results are unaffected by these factors, as reported in a previous preliminary analysis [40]. However, there may be other specific conditions and treatments we have not yet examined, that could affect SeptiCyte RAPID performance.

Although the dynamic range of SeptiCyte RAPID is broad and extends well below and above the 4,000-11,000 WBC/uL normal reference range, we have not yet completed an evaluation of the assay on severely neutropenic patients, although in a previous preliminary analysis we reported that SeptiCyte performance was maintained in patients treated with anti-neoplastic drugs [40]. It is possible that SeptiCyte RAPID scores might be skewed by selective leukopenias due to disease or medications, for example T cell depletion in HIV / AIDS. However, it is known from single cell sequencing studies that both PLAC8 and PLA2G7 are expressed across a range of different white cell types [41] which would mitigate the effect of a selective leukopenia.

The recently updated Surviving Sepsis Campaign (SSC) guidelines (2021) divide patients into three groups (low, intermediate, and high) based on sepsis probability, and recommend appropriate evaluation and treatment of these groups [42]. SeptiCyte RAPID aligns well with these guidelines and could have a role in supporting their implementation. The SSC ‘low sepsis probability’ group has, under previous guidelines, been treated early resulting in poor antibiotic stewardship. The new guidelines recommend deferring antibiotics, and monitoring and evaluate for other etiologies that may underlie the presenting symptoms. A SeptiScore <5 (Band 1) with a sensitivity of 0.94 in this group of patients could potentially support the deferring of antibiotics, at least until clinical microbiology results became available, thereby facilitating antibiotic and diagnostic stewardship. For patients falling in the SSC ‘intermediate sepsis probability (without shock)’ group, the SSC recommendation is a rapid assessment of infectious versus non-infectious cause of the illness. In this probability range, the information typically obtained from other clinical variables could be augmented by information from SeptiCyte RAPID to shift the post-test probability of sepsis to either a lower or higher value (Figure 4). Per the SSC guidelines, those patients in the ‘high sepsis probability’ group, differentiated by the presence of shock, should appropriately be treated within 1 hour of recognition. SeptiScores in Band 4 (>7.5) in this patient group, with specificity 0.9 or greater, would provide evidence for a high probability of sepsis and the continuation of antibiotics.

Although SeptiCyte RAPID provides a fast turnaround time (∼1 hour) between pipetting a blood sample into the SeptiCyte RAPID cartridge and generating a test report, by the time patient blood is drawn and delivered e.g. to a STAT lab, the total time between blood draw and presentation of the test result to an attending clinician is likely to be 1.5-2 hours. Although this is longer than requirements to implement a 1-hour sepsis bundle, SeptiCyte RAPID would provide timely information with respect to implementing a 3-hour sepsis bundle, which has been shown to have a low level of compliance in US hospitals [43,44]. Compliance with a 3-hour sepsis bundle in patients with severe sepsis and septic shock has been shown to improve survival and reduce overall costs [44].

## 5. Conclusions

This study validates SeptiCyte RAPID for discrimation of sepsis from non-infectious systemic inflammation, and for the estimation of probability of sepsis on the first day of ICU admission, under either the Sepsis-2 or Sepsis-3 frameworks. With a turnaround time of ∼1 hour, SeptiCyte RAPID provides actionable test results within a clinically relevant timeframe. SeptiCyte RAPID may thus provide clinical utility through helping to guide patient management decisions on the implementation and timing of sepsis bundles.

## Supplementary Materials

The following supporting information can be downloaded at: www.mdpi.com/xxx/ Balk et al. (2023) Supplementary Information

## Author Contributions

Conceptualization: RFD, SC, RBB, DS, TDY, JTK; Methodology: RFD, SC, DS, TDY, KN, JTK; Investigation: RB, AME, GSM, RRM III, BKL, JPB, ML, SO, RER, FRDA, VKS, NRA, JAG, MY, GP, EG, MA, JPP, JAK, TvdP, MJS, BPS, PMCKK, JL, EB, SK, JTK; Supervision: SC, RFD, RB, AME, GSM, RRM III, BKL,TvdP, MJS, JL, EG; Data Curation & Review: SC, DS, KN, JTK; Clinical Validation (Reference Diagnoses): SO, RER, ML; Custom software: DS, KN; Visualization: DS, KN, TDY; Formal analysis: DS, KN, TDY, SC; Writing: original & successive drafts: TDY, RBB, RFD; Writing: review and editing: TDY, RBB, RFD, SC, JTK, RB, GSM, JAK, BPS, TvdP, PMCKK; Project Administration: TDY, SC.

## Institutional Review Board Statement

Ethics approval for the MARS trial was given by the medical ethics committee of AMC, Amsterdam (approval # 10-056C). Ethics approvals for the VENUS trial were given by the relevant Institutional Review Boards as follows: Intermountain Medical Center/Latter Day Saints Hospital (approval # 1024931); Johns Hopkins Hospital (approval # IRB00087839); Rush University Medical Center (approval # 15111104-IRB01); Loyola University Medical Center (approval # 208291); Northwell Healthcare (approval #16-02-42-03). Ethics approvals for the NEPTUNE trial were given by the relevant Institutional Review Boards as follows: Emory University (approval # IRB00115400); Grady Memorial Hospital (approval # 00-115400); Rush University Medical Center (approval # 19101603-IRB01); University of Southern California Medical Center (approval # HS-19-0884-CR001). All methods used in this study were carried out in accordance with the relevant guidelines and regulations.

## Informed Consent Statement

All subjects, or their legally authorized representatives, gave informed consent for participation in this study.

## Data Availability Statement

The datasets used and/or analysed during the current study are available from the corresponding author upon reasonable request.

## Acknowledgments

The authors thank the clinical study coordinators of the NEPTUNE study: Joyce D. Brown (Rush University), Liliacna Jara (University of Southern California), Leona Wells and Maya C. Whaley (Grady Memorial Hospital). The authors also thank the laboratory technical staff at each institution,at Reach Bio (Seattle WA), and at Biocartis (Mechelen, Belgium).

