## Supplementary Information for "Validation of SeptiCyte RAPID to discriminate sepsis from non-infectious systemic inflammation"

### Table of Contents

1. Study Cohorts
2. Definitions of SIRS, Sepsis, and Comparator (Reference Method)
3. Imputation of Missing Clinical Data Values
4. Data Subsets and Statistical Methods for Exploratory Data Analysis
5. Multivariable Analysis
6. SeptiCyte RAPID - SeptiCyte LAB correlation
7. Gene Expression Signature: Biological Roles of PLA2G7 and PLAC8
8. Sepsis Cases Stratified by Culture Results
9. Sepsis Cases Stratified by Pathogen Type
10. The Relation Between SeptiCyte RAPID and Organ Dysfunction
11. References

### 1. Study Cohorts

Clinical validation of SeptiCyte RAPID used PAXgene blood RNA samples from retrospective (N=356) and prospective (N=63) patient cohorts, as described in the main text. A flow diagram describing the origin of all samples used in the study is provided in **Figure S1**.

**Figure S1.** Diagram describing origin of samples used in the present study. Abbreviation: VENUS ext., extension of the VENUS Trial.

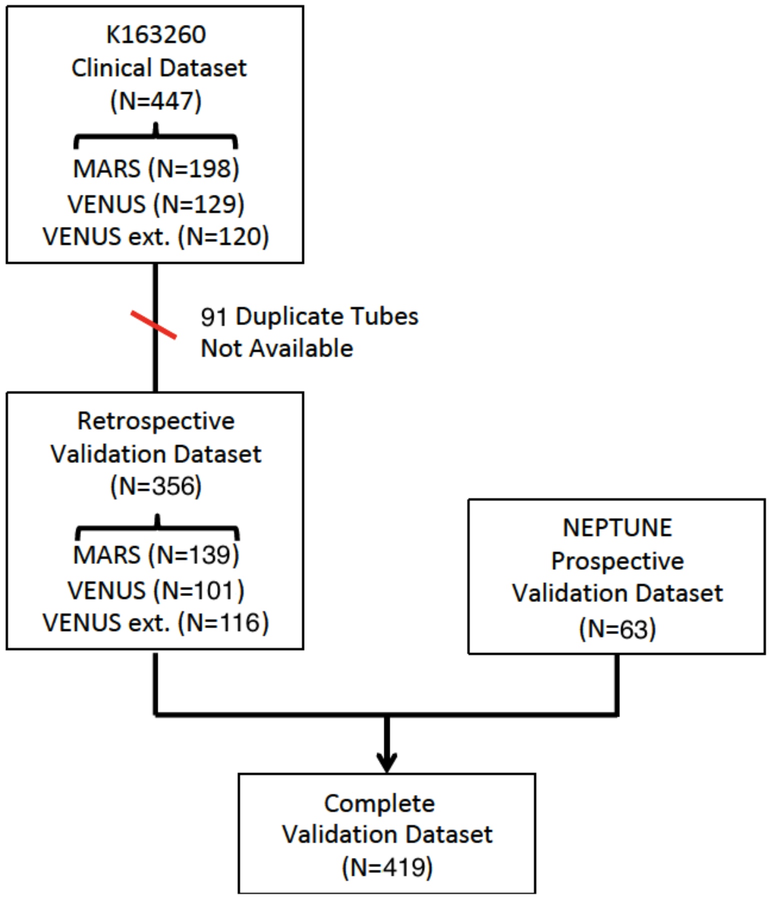

### 2. Definitions of SIRS, Sepsis, and Comparator (Reference Method)

Although sepsis has been known as a medical condition since antiquity, the modern understanding of sepsis begins in the 1990s and has evolved through three phases, each characterized by a different definition (Gul et al., 2017). Interwoven through this evolution is also a consideration of SIRS, the systemic inflammatory response syndrome.

*SIRS* (Balk, 2014) - The definition of SIRS arose an American College of Chest Physicians/Society of Critical Care Medicine-sponsored 'sepsis definitions conference' held in Chicago, IL in August 1991 (Bone et al., 1992). This was the same conference that gave rise to the Sepsis-1 definition (below). SIRS, which occurs in response to various infectious and non-infectious causes, was defined to consist of one or more of the following factors: (1) temperature >38°C or <36°C; (2) heart rate >90 beats per minute; (3) respiratory rate >20 breaths per minute (BPM) or $PaCO_2$ <32 mmHg; (4) white blood cell count >12,000/mm$^3$, <4,000/mm$^3$, or >10% immature (band) forms. Motivation for defining SIRS in this relatively non-specific way came from view that it would focus attention toward developing effective strategies to limit excessive inflammatory responses in the patient, due to various underlying causes (sepsis, trauma, burns, pancreatitis, etc.) Criticism of the SIRS definition has focused on the lack of specificity that this definition entails (Vincent et al., 2013).

*Sepsis-1* (Bone et al., 1992) - This definition arose from the same 'sepsis definitions conference' referenced above. Sepsis was defined as the condition occurring when 2 or more SIRS criteria are met as a consequence of infection. Severe sepsis was defined as sepsis with associated organ dysfunction. Septic shock was defined as sepsis-induced hypotension.

*Sepsis-2* (Levy et al., 2003) - documented or suspected infection, together with 2 or more SIRS-like criteria being met: temperature > 38.3°C or <36°C; heart rate >90 BPM or >2 SD above the normal value for age; respiratory rate >30 BPM; altered mental status; significant edema or positive fluid balance; hyperglycemia in the absence of diabetes. More detailed criteria for organ dysfunction were specified. The Sepsis-2 definition could be considered a refinement and elaboration of the Sepsis-1 definition, which served as the basis for development of treatment guidelines (Levy et al., 2010).

<u>*Sepsis-3*</u> (Singer et al., 2016) - life-threatening organ dysfunction caused by a dysregulated host response to infection. In turn, organ dysfunction is defined as an acute increase in total Sequential Organ Failure Assessment (SOFA) score by 2 or points, as a consequence of the infection. The Sepsis-3 definition is a fairly major departure from Sepsis-1/2 in that it does not use the term ‘SIRS’, and emphasizes organ dysfunction. It also does not distinguish between sepsis and severe sepsis.

Another point to recognize is that, with the emphasis on organ dysfunction, a clinician operating strictly under the Sepsis-3 definition could potentially miss the early warning signs of sepsis before organ dysfunction was obvious. This point of concern has been raised by a number of authors (Zhang et al., 2016; Tusgul et al., 2017; Dorsett et al., 2017; Sartelli et al., 2018; Kim & Park, 2019; Lukaszewski et al., 2022), and is relevant to our own study.

Our study, which can be viewed as an extension of the previous study of Miller et al. (2018), was based on definitions of SIRS and Sepsis-2 given above. This was considered appropriate, as 240/419 (57.2%) of the samples in the present study were drawn from patients recruited in trials that predated publication of the Sepsis-3 definition: 139 subjects from the MARS trial between June 2013 and November 2013; and 101 from the VENUS trial between May 2014 and August 2015. These recruitment periods occurred before publication of the Sepsis-3 definition, which occurred on February 23, 2016 (Singer et al., 2016). An additional 116 subjects were recruited in the VENUS Supplement trial from March to August 2016, under a protocol that had been approved by the relevant Institutional Review Boards before the publication date of Singer et al. (2016). An additional 63 subjects were recruited in the NEPTUNE trial from May 2020 - April 2021. It was decided to recruit in these later trials in alignment with the SIRS and Sepsis-2 definitions, to maintain consistency with the previous MARS and VENUS recruitments.

A further consideration was that organ dysfunction (the focus of Sepsis-3) occurs at the phenotypic level, as modulated by protein structure & function, whereas SeptiCyte

RAPID is a gene expression-based assay. Changes in gene expression occur some hours before changes in protein expression occur. By this logic, the Sepsis-2 definition would appear more appropriate than the Sepsis-3 definition for our study. A similar argument has been made by Lukaszewski et al. (2022), in support of studies investigating an independently discovered sepsis signature.

To define the "Ground Truth" for our study, an external three-member panel of experienced physicians not involved in the clinical care of the patients performed a three-way patient classification (sepsis, SIRS or indeterminate) by chart review in accordance with the SIRS and Sepsis-2 definitions (Levy et al., 2003). This approach follows the general framework defined by Klein-Klouwenberg and colleagues for employing an expert panel for assigning infection probability estimates in studies on sepsis (Klein-Klouwenberg et al., 2012; 2013; 2015; Lopansri et al., 2019). Three algorithms for this Retrospective Physician Diagnosis (RPD) process were used:

*Consensus:* Two or three RPD panelists agreed that a patient had either sepsis or SIRS. Indeterminates, which occurred when two or three panelists made a call of 'indeterminate' or when all three panelists disagreed, were excluded.

*Unanimous:* All three RPD panelists agreed that a patient had either sepsis or SIRS. (A unanimous call of indeterminate was also theoretically possible.)

*Forced:* When a patient was initially called "indeterminate" by the RPD panelists, the panelists were then forced to make a consensus or unanimous call of sepsis or SIRS.

The RPD panelists received a data package for each patient, consisting of 39 data elements as specified in **Table S1**.

**Table S1.** Clinical data elements made available to the RPD panelists (per patient).

| No. | Description |
|---|---|
| **1. DEMOGRAPHICS** | |
| 1 | Age of subject |
| 2 | Race of subject |
| 3 | Sex of subject |
| 4 | Source of patient, pre-ICU admission |
| **2. VITAL SIGNS** | |
| 5 | Minimum mean arterial blood pressure (MAP min) |
| 6 | Maximum mean arterial blood pressure (MAP max) |
| 7 | Minimum measured body temperature |
| 8 | Maximum measured body temperature |
| 9 | Minimum measured heart rate |
| 10 | Maximum measured heart rate |
| **3. CLINICAL LAB PARAMETERS** | |
| 11 | Minimum white blood cell (WBC) count |
| 12 | Maximum white blood cell (WBC) count |
| 13 | pH of the blood |
| 14 | Maximum measured blood glucose concentration |
| 15 | Minimum measured blood glucose concentration |
| 16 | Concentration of serum lactate |
| 17 | Concentration of C-reactive protein (CRP) |
| **4. CALCULATED QUANTITIES** | |
| 18 | Number of SIRS criteria observed at enrollment |
| 19 | APACHE (Acute Physiologic Assessment and Chronic Health Evaluation) score, if available |
| 20 | SOFA (Sequential Organ Failure Assessment) score, if available |
| 21 | SOFA respiratory component (based on PaO2/FiO2 ratio) |
| 22 | SOFA coagulation component (based on average Platelet count) |

| No. | Description |
|---|---|
| 23 | SOFA liver component (based on Serum bilirubin level) |
| 24 | SOFA cardiovascular component (based on Mean arterial blood pressure) |
| 25 | SOFA central nervous system component (Glasgow Coma Scale score) |
| 26 | SOFA renal component<br>(based on serum creatinine value or urine output per day) |
| **5. INFECTION PARAMETERS** | |
| 27 | Consensus of the Site Principal Investigators' impression of infection status (none, possible, probable or definite) in the first 24 hours after ICU admission. (This assessment is made retrospectively, upon discharge from ICU.) |
| 28 | Microorganisms identified in microbiological tests |
| 29 | Whether antimicrobials were administered during ICU stay (Y/N) |
| 30 | Antimicrobial treatments administered during ICU stay |
| 31 | Occurrence of a new infection within 7 days of ICU admission<br>(i.e. an infection that was not present at ICU admission) (Y/N) |
| 32 | Location of new infection, if apparent |
| 33 | Microorganism responsible for the new infection |
| **6. OTHER DATA** | |
| 34 | Results of imaging procedures (e.g. chest X-ray, CT scan, pelvic CT scan) |
| 35 | Site Principal Investigators' comments (verbatim transcription of text from Site Principal Investigators, if added during discharge assessment process) |
| 36 | Whether subject required mechanical ventilation (Y/N) |
| 37 | ICU length of stay (days) |
| 38 | Hospital length of stay (days) |
| 39 | Death during hospitalization (Y/N) |

### 3. Imputation of Missing Clinical Data Values

For the individual clinical laboratory variables, data were collected over the 24-hour period following ICU admission. Missing values were imputed using the multiple imputation algorithm Amelia (Honaker et al., 2011) (**Table S2**). Multiple imputation reduces bias and increases accuracy over point-imputation methods such as “mean” or “median” imputation. Amelia produces $n_{imputed}$ datasets which are then processed with downstream algorithms as needed. In this case, $n_{Imputations}$ was set to 5.

Note: the #(SIRS criteria observed at enrollment) was calculated on the basis of clinical data recorded over the 24 hour study enrollment period. This quantity is calculated as a sum with 1 point for each of the following:

- Heart Rate > 90 beats / minute
- Respiratory Rate > 20 breaths / minute
- Core temperature < 36.0 °C or > 38.0 °C
- White Blood Cell count < 4,000/uL or >11,000/uL or % Bands > 10%

In order for a patient to be enrolled in the study, calculation of the #(SIRS criteria at enrollment) was necessary. Therefore, there are no missing values for this parameter.

The 24-hour study enrollment period precedes and partially overlaps the first 24 hours after ICU admission. Thus, although the #(SIRS criteria observed at enrollment) must have been recorded for the patient to have been enrolled in the study, data for some of the clinical laboratory variables may not have been not recorded during the first 24 hours after ICU admission, leading to missing values in the clinical line data file.

**Table S2.** Imputation of missing values in the clinical line data, by means of the Amelia software package.

| **Variable** | **# values measured** | **# values imputed** | **Imputed global median** | **Units** |
|---|---|---|---|---|
| CRP.Level | 46 | 373 | 10.5 | mg/L |
| Bands_Percent.Max | 62 | 357 | 1.25 | percent |
| Bands_Percent.Min | 63 | 356 | 1.10 | percent |

| Variable | # values measured | # values imputed | Imputed global median | Units |
|---|---|---|---|---|
| Platelets.Max | 156 | 263 | 199.5 | $x10^3$/uL |
| Lactate.Level | 279 | 140 | 2.2 | mmol/L |
| SOFA score | 290 | 129 | 6 | unitless |
| Respiratory.Rate.Min | 290 | 129 | 16.5 | breaths/min |
| Respiratory.Rate.Max | 285 | 124 | 26 | breaths/min |
| Platelets.Min | 297 | 122 | 182 | $x10^3$/uL |
| Mean.Art.Pressure.Max | 324 | 95 | 110.5 | mm Hg |
| $\log_2$ PCT | 347 | 72 | -0.34 | $\log_2$ (ng/mL) |
| Temp.Min | 355 | 64 | 36.1 | °C |
| Temp.Max | 366 | 53 | 37.4 | °C |
| Glucose.Max | 390 | 29 | 165 | mg/dL |
| Glucose.Min | 393 | 26 | 122.5 | mg/dL |
| WBC.Max | 403 | 16 | 13.7 | $x10^3$/uL |
| WBC.Min | 407 | 12 | 10.78 | $x10^3$/uL |
| Mean.Art.Pressure.Min | 401 | 9 | 63 | mm Hg |
| HeartRate.Min | 417 | 2 | 74 | beats/min |
| HeartRate.Max | 419 | 0 | -- | beats/min |
| Age | 419 | 0 | -- | years |
| SeptiScore | 419 | 0 | -- | unitless |

### 4. Data Subsets and Statistical Methods for Exploratory Data Analysis

Different subsets of the line data file were considered, depending upon the particular analysis being done (**Table S3**).

**Table S3.** Utilization of clinical variables in downstream analyses. Single variable performance (AUC) was evaluated for the sepsis vs. SIRS comparison, with no values imputed, and with consensus RPD as reference. Fig. 4 refers to a figure in the main text. ND = not determined (categorical variable).

| RPD element No. | Description | Single variable performance AUC (N) | Used in multivariable analysis (Fig. 4)? |
|---|---|---|---|
| | **1. DEMOGRAPHICS** | | |
| 1 | Age of subject | 0.58 (378) | yes |
| 2 | Race of subject | ND | yes (African-American or not) |
| 3 | Sex of subject | ND | yes |
| | **2. VITAL SIGNS** | | |
| 5 | Minimum mean arterial blood pressure (MAP min) | 0.62 (370) | no |
| 6 | Maximum mean arterial blood pressure (MAP max) | 0.60 (295) | yes |
| 7 | Minimum measured body temperature | 0.53 (320) | yes |
| 8 | Maximum measured body temperature | 0.67 (328) | yes |
| 9 | Minimum measured heart rate | 0.57 (377) | yes |
| 10 | Maximum measured heart rate | 0.60 (378) | yes |
| -- | Respiratory rate .Min | 0.61 (262) | no* |
| -- | Respiratory rate .Max | 0.63 (265) | no* |
| | **3. CLINICAL LAB PARAMETERS** | | |
| 11 | Minimum white blood cell (WBC) count | 0.54 (369) | yes |
| 12 | Maximum white blood cell (WBC) count | 0.56 364) | yes |
| -- | Platelets.Max | 0.55 (140) | no* |
| -- | Platelets.Min | 0.52 (269) | no* |
| -- | Bands_Percent.Max | 0.77 (57) | no* |
| -- | Bands_Percent.Min | 0.78 (58) | no* |
| 14 | Maximum measured blood glucose concentration | 0.55 (351) | yes |
| 15 | Minimum measured blood glucose | 0.52 (355) | no** |

| RPD element No. | Description | Single variable performance AUC (N) | Used in multivariable analysis (Fig. 4)? |
|---|---|---|---|
| | concentration | | |
| 16 | Concentration of serum lactate | 0.54 (253) | yes |
| 17 | Concentration of C-reactive protein (CRP) | 0.80 (41) | no* |
| -- | Concentration of Procalcitonin (PCT) | 0.81 (310) | yes |
| -- | SeptiScore (0-15) | 0.85 (378) | yes |
| | **4. CALCULATED QUANTITIES** | | |
| 18 | Number of SIRS criteria observed | 0.67 (378) | yes |
| 19 | APACHE (Acute Physiologic Assessment) score Chronic Health Evaluation) score, if available | 0.60 (376) | no |
| 20 | SOFA (Sequential Organ Failure Assessment) score, if available | 0.60 (264) | no* |

* not included in the multivariable analysis, because of too many missing values (>30%)

### 5. Multivariable Analysis

A custom R script was written to evaluate the classification performance of all 32,767 possible combinations of SeptiScore with any of the 14 clinical variables specified in column 4 of **Table S3**. AUC was used as performance measure with Consensus RPD as comparator.

The algorithm used for multivariable analysis consists of the following steps:

Step 1. As indicated in **Table S3**, 13 of the 39 variables used in the RPD process were employed in the multivariable analysis, as they satisfied the following criteria:

1) routinely used as a diagnostic aid in the differential diagnoses of sepsis;
2) routinely available in the first 24 hours after ICU admission;
3) no more than 30% missing values requiring imputation.

Procalcitonin and SeptiScore were also used, making 15 variables in all.

Step 2. The data were partially incomplete. Missing values were imputed with Amelia, a multiple imputation algorithm (Honaker et al., 2011) that produces $n_{imputed}$ datasets, which can then be processed with downstream algorithms as needed. It has been argued that multiple imputation reduces bias and increases accuracy over point-imputation methods such as "mean" or "median" imputation (Honaker et al., 2011). In this case, $n_{Imputations}$ was set to 5.

Step 3. From the pool of 15 variables listed in Table S3, all possible unordered subsets of variables were defined, with no repetitions. Each such combination consisted of 1 to 15 variables. The total number of possible combinations is given by the following formula:

$$\sum_{j=1}^{15} \binom{15}{j} = \sum_{j=1}^{15} \frac{15!}{(j!)(15-j)!} = 32{,}767 \quad \text{(Eq. S1)}$$

Step 4. For each possible combination of variables, a multivariable model was constructed by logistic regression, and the AUC was calculated with consensus RPD as the comparator.

Step 5. The distribution of AUC for all models was plotted (Figure 5 in main text) by dividing into three histograms for (i) models not containing SeptiScore or PCT, (ii) models containing PCT but not SeptiScore, and (iii) models containing SeptiScore (which may also include PCT).

### 6. SeptiCyte RAPID - SeptiCyte LAB Correlation

**Figure S2.** Correlation plot for SeptiCyte LAB vs. SeptiCyte RAPID. The scores for SeptiCyte RAPID (2 gene assay, x-axis scale 0-15) and SeptiCyte LAB (4 gene assay, x-axis scale 0-10) were measured for 356 patients from retrospective clinical trials performed in the USA (VENUS study) and Europe (MARS study). A least-squares linear fit to the data points (y = -0.48 + 0.82x) had Pearson's sample correlation coefficient r = 0.88.

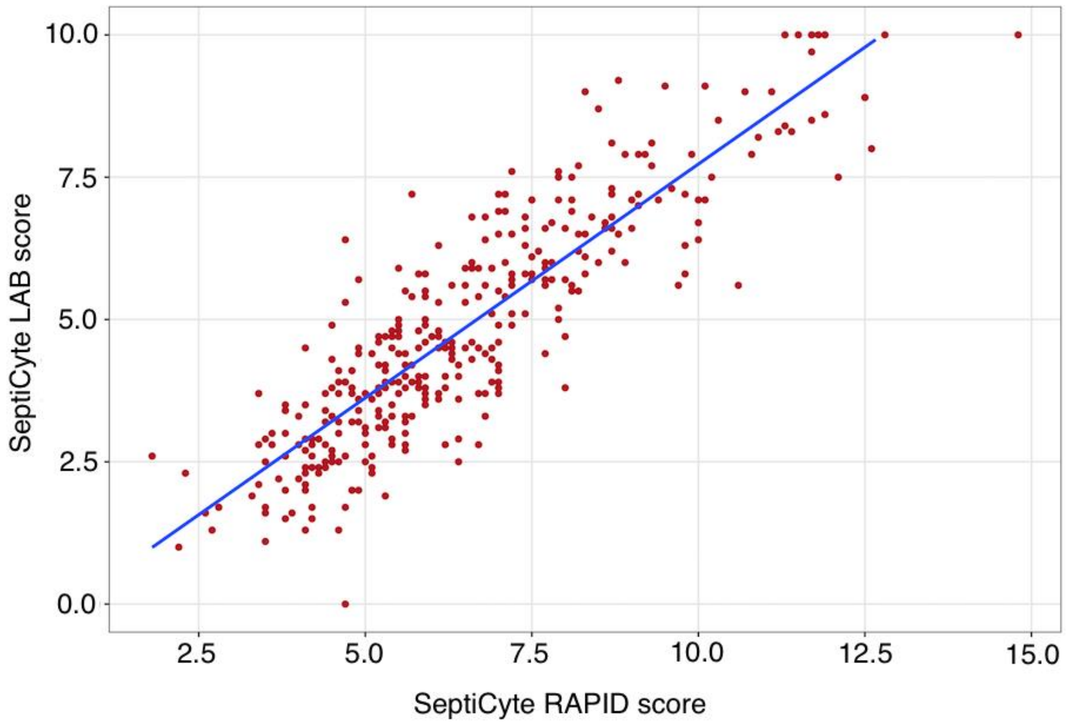

### 7. Gene Expression Signature: Biological Roles of PLA2G7 and PLAC8

The SeptiCyte RAPID signature measures the relative expression levels of two immune-related mRNAs, PLAC8 (Placenta-associated 8) and PLA2G7 (Phospholipase A2 Group 7). The signature was discovered using a purely bioinformatic approach, as described previously (McHugh et al., 2015).

**PLAC8** is reportedly an interferon inducible gene (Pankla et al., 2009) and is expressed in a variety of immune cells (spleen, lymph nodes), including plasmacytoid dendritic cells. It has putative roles in the optimal function of neutrophils following the uptake of bacteria (Ledford et al., 2007), the clearance of Chlamydia (Johnson et al., 2012, 2013), and the host response to viral infections (Wieland et al., 2004). Related to this function is that it is expressed highly in neutrophil azurophilic granules (Velasquez et al., 2022). In sepsis, it is up-regulated across a broad range of different peripheral blood cell types including plasmacytoid dendritic and natural killer cells (Reyes et al., 2020).

**PLA2G7** encodes the protein platelet-activating factor (PAF) acetylhydrolase, a secreted enzyme primarily produced by macrophages. This enzyme catalyzes the degradation of PAF and hydrolyses the oxidized short chain phospholipids of low-density lipoproteins (LDL), thereby releasing pro-inflammatory mediators (lysophospholipids and oxidized fatty acids). High plasma levels have been found to correlate with sepsis survival (Huang, 2018) and decreased levels have been found in sepsis (Gomes et al., 2006; Yang et al., 2010; Huang, 2020).

### 8. Sepsis Cases Stratified by Culture Results

**Figure S3** presents a stratification of patients with respect to both the sepsis/SIRS assessment by consensus RPD (with 41 indeterminates not counted), and also whether pathogen identification by culture or PCR was obtained or not. Of the 154 patients called septic by consensus RPD, 31 did not have positive culture or PCR results, but were still discriminated significantly away from SIRS with AUC = 0.75 ($p = 4.82 \times 10^{-5}$). It is perhaps unsurprising that culture/PCR-positive sepsis cases would tend toward higher AUC values than culture/PCR-negative sepsis cases. Likely explanations would include: (1) that culture-positive sepsis exhibits a more pronounced inflammatory response to presence of more ‘pathogen-associated molecular pattern molecules’ (PAMPs); and (2) that positive culture results would influence the RPD decision process more towards a sepsis diagnosis.

**Figure S3.** Stratification of patient cohort according to diagnosis by consensus RPD, and by culture or PCR findings. The first dimension is diagnosis of sepsis or SIRS by consensus RPD (with indeterminates excluded). The second dimension is whether positive or negative results were returned by microbiological culture for bacteria or fungi, or by PCR (or RT-PCR) for viruses.

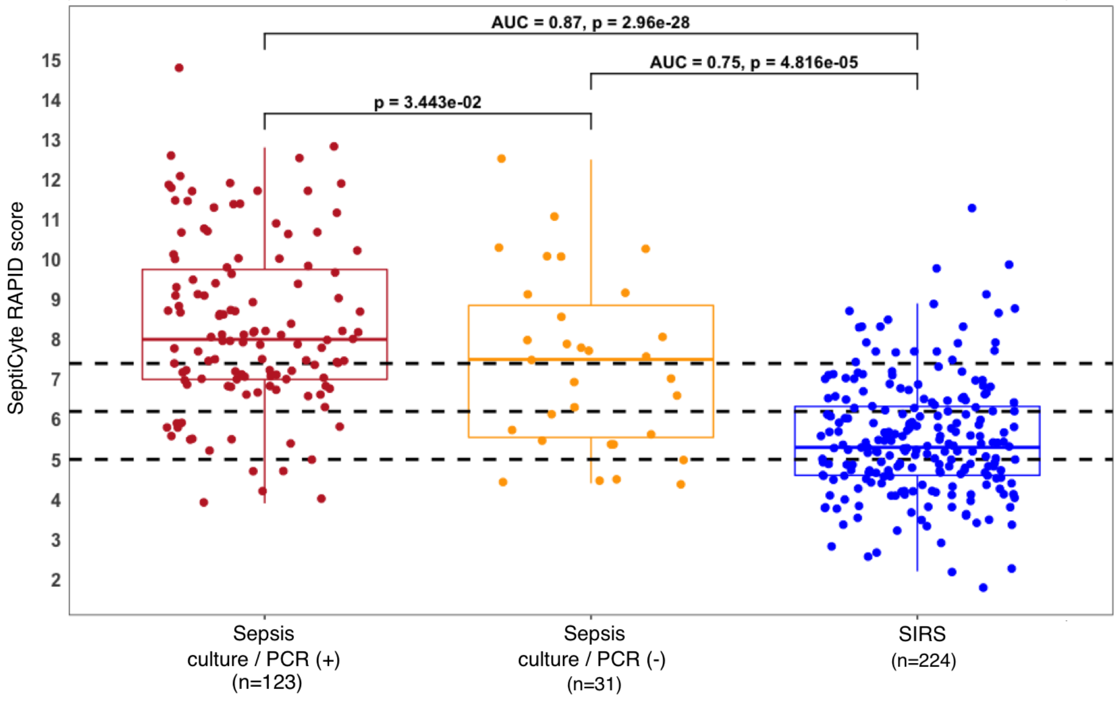

### 9. Sepsis Cases Stratified by Pathogen Type

Out of 419 ICU patients, there were 154 (36.8%) sepsis cases according to Consensus RPD. Of these, 100 (64.9%) were considered to have one or more identifiable pathogens (bacterial, fungal, viral) as the underlying causative agent(s). **Figure S4** represents the distribution of identified pathogens by means of pie charts, with panel (A) at the level of bacterial, viral, fungal pathogens and panel (C) at the level of bacterial genera and species. The remaining panels (B) and (D) show the SeptiScore distributions for patients with bacterial vs. mixed (bacterial/viral/fungal) infections (Panel B), and for patients with Gram (-) vs. Gram (+) bacterial infections (Panel D). No significant differences in SeptiScore distributions were found in the comparisons in either panel. Thus, in the cohorts studied here, SeptiScore does not appear to be significantly affected by the type of pathogen underlying a sepsis event.

**Figure S4.** Septic patients stratified by pathogen type and bacterial species. Sepsis diagnosis was by consensus RPD. Only those septic patients with pathogens identified by culture or PCR are represented (N=100). (A) Pie chart for pathogens by type (bacterial, viral, fungal). (B) Cumulative SeptiScore distributions for patients with identified bacteria (N=89) vs. acute viral, fungal or mixed infections (N=13). A Kolmogorov-Smirnov test indicates no significant difference (p=0.375). (C) Pie chart for bacteria by species. (D) Cumulative SeptiScore distributions for patients with identified Gram positive (N=55) vs. Gram negative (N=36) infections. A Kolmogorov-Smirnov test indicates no significant difference (p=0.264). Pie charts were drawn with the R package ggplot2.

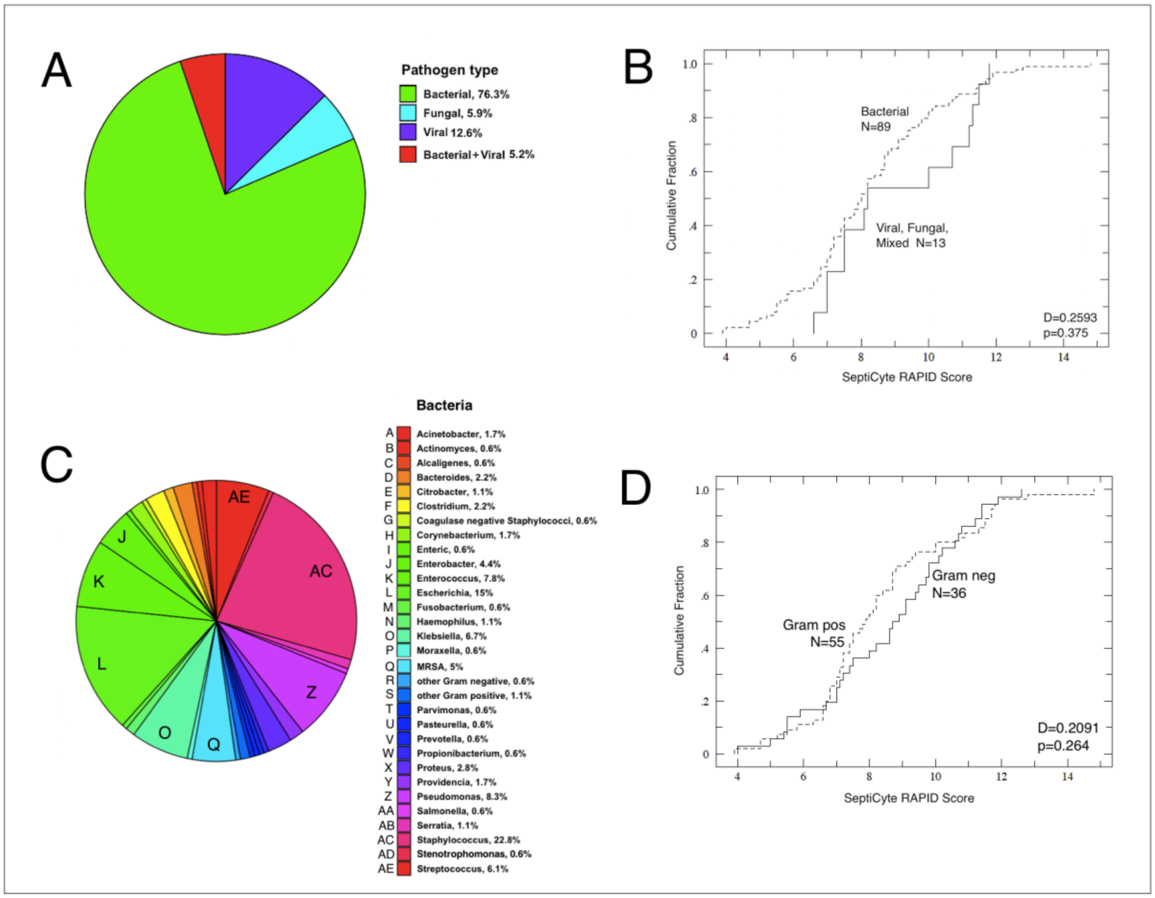

The observed distributions of the most commonly occurring pathogen types found in sepsis-associated positive cultures (not restricted to blood cultures) are listed **Table S4.** For context, this table also shows the corresponding frequencies observed in blood cultures from hospitalized US patients (taken from a large survey study of 82,569 bacterial blood culture isolates reported to The Surveillance Network (TSN) Database-USA by 268 laboratories; Karlowsky et al., 2004).

**Table S4.** Breakdown of specific pathogen genera and species (present at >1%), isolated in cultures from septic patients (N=100 patients total). Sepsis diagnosed by consensus RPD.

| Pathogen | Observed in sepsis cases, present study* | Karlowsky et al. (2004) blood culture |
|---|---|---|
| Gram (+) only | 31 (31%) | -- |
| Gram (-) only | 30 (30%) | -- |
| Gram (+/-) (both) | 26 (26%) | -- |
| Bacterial/viral | 8 (8%) | -- |
| Bacterial/fungal | 4 (4%) | -- |
| Staphylococcus aureus | 34 (including 9 MRSA) (34%) | 16.5 % |
| Escherichia coli | 25 (25%) | 7.2 % |
| Pseudomonas aeruginosa | 12 (12%) | 2.5 % |
| Klebsiella spp. | 9 (9%) | 3.6 % |
| Enterococcus faecalis | 8 (8%) | 8.3 % |
| Proteus spp. | 5 (5%) | 0.9 % |
| Bacteroides fragilis | 3 (3%) | < 0.2% |
| Enterococcus faecium | 3 (3%) | 3.5 % |
| Candida spp. | 3 (3%) | < 0.2% |
| Streptococcus pneumoniae | 2 (2%) | 2.3 % |
| Clostridium dificile | 2 (2%) | < 0.2% |
| Clostridium perfringens | 2 (2%) | < 0.2% |
| Corynebacterium spp. | 2 (2%) | < 0.2% |
| Citrobacter freundii | 2 (2%) | 0.3 % |
| *excludes cases where positive culture result occurred > 5 days after ICU admission (considered not relevant to the admitting sepsis event), or where obvious contaminant | | |

There were six SARS-CoV-2 positive patients in the prospective cohort, five of whom were called septic and one SIRS by consensus RPD. In Figure 1B (main text), the prospective cohort ROC curves for forced (black), consensus (blue) and unanimous (red) RPD, had AUC = 0.86, 0.90 and 0.95 respectively. When the six COVID-19 patients were removed from the prospective cohort and the AUC analyses repeated, no significant effect was found (AUC = 0.85, 0.90, 0.98 for forced, consensus, unanimous RPD respectively).

**10. SeptiCyte RAPID Appears Independent of Organ Dysfunction**

In our previous paper on the 4-gene SeptiCyte LAB signature (McHugh et al., 2015) we asked whether organ dysfunction was a potential confounder of the SeptiScore. The conclusion we reached (Figure 5 therein) was that “with respect to discrimination of cases from controls by SeptiCyte Lab, any confounding effect of disease severity, as measured by APACHE or SOFA scores, appeared small”.

We conducted a similar analysis, in which patients’ SeptiCyte RAPID scores were stratified by SOFA. The dependence of procalcitonin (PCT) on SOFA was also examined for comparison. The results of this new analysis are shown in **Figure S5**. Similarly to SeptiCyte LAB we see little, if any, dependence of the RAPID SeptiScore on SOFA score. We interpret this to mean that SeptiCyte RAPID is not measuring or responding to organ dysfunction, but rather to some other characteristic of the septic process, possibly occurring earlier than organ dysfunction. A similar conclusion was reached for an independently discovered gene expression signature proposed for sepsis diagnosis (Lukaszewski et al., 2022).

**Figure S5.** RAPID SeptiScore as function of SOFA score. (A, B) RAPID SeptiScores and procalcitonin (PCT) values, respectively, for sepsis (red) vs. SIRS (gray) groups, parsed into different SOFA bins. Forced RPD was used for the sepsis vs. SIRS assignments, and only those patients having both SeptiScores and PCT values were used. (C) SeptiScore AUC for the sepsis vs. SIRS discrimination, as a function of the SOFA sliding window position.

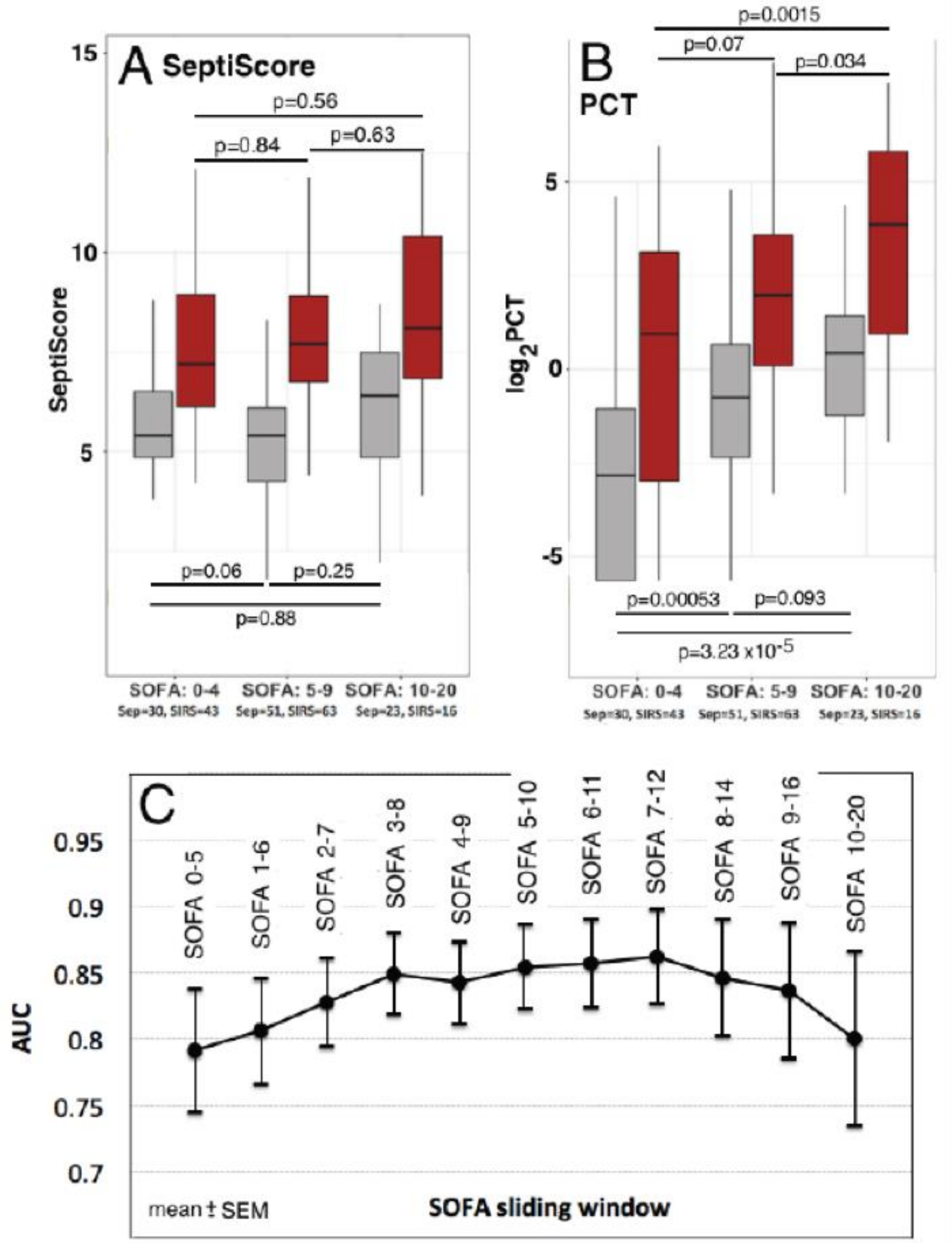
A SeptiScore
SeptiScore
p=0.56
p=0.84
p=0.63
p=0.06
p=0.25
p=0.88
B PCT
$log_2$PCT
p=0.0015
p=0.07
p=0.034
p=0.00053
p=0.093
p=3.23 x10$^{-5}$
SOFA: 0-4
Sep=30, SIRS=43
SOFA: 5-9
Sep=51, SIRS=63
SOFA: 10-20
Sep=23, SIRS=16
C
AUC
SOFA 0-5
SOFA 1-6
SOFA 2-7
SOFA 3-8
SOFA 4-9
SOFA 5-10
SOFA 6-11
SOFA 7-12
SOFA 8-14
SOFA 9-16
SOFA 10-20
mean ± SEM
SOFA sliding window
